# Time-Varying Cardiovascular Risk of Febuxostat versus Allopurinol in Gout: A One-stage Meta-analysis

**DOI:** 10.64898/2025.12.29.25343180

**Authors:** Qige Wei, Yang Shi, Rui Hu, Hui Wang, Yan Li, Li Luo, Dan Shao, Jianglin Zhao, Shengzhao Zhang

**Affiliations:** Department of Pharmacy, Central Hospital of Karamay, Karamay, China; Department of Pharmacy, Karamay Hospital of People’s Hospital of Xinjiang Uygur Autonomous Region, Karamay, China; Department of Pharmacy, Karamay Municipal Hospital of Integrated Traditional Chinese and Western Medicine (Karamay Municipal People’s Hospital), Karamay, China; Department of Cardiology, Central Hospital of Karamay, Karamay, China; Department of Cardiology, Karamay Hospital of People’s Hospital of Xinjiang Uygur Autonomous Region, Karamay, China

**Keywords:** Gout, Febuxostat, Allopurinol, Cardiovascular Events, All-cause Mortality

## Abstract

**Objectives:** Whether febuxostat is associated with an increased cardiovascular risk compared to allopurinol in patients with gout remains controversial, with major randomized trials reporting conflicting results. This study aims to perform a comprehensive meta-analysis using reconstructed individual participant data (IPD) to evaluate the time-varying cardiovascular and mortality risk of febuxostat versus allopurinol in gout.

**Methods:** We conducted a one-stage individual participant data meta-analysis. PubMed, Embase, and Cochrane Library were searched up to November 2025 for randomized controlled trials and propensity score-matched observational studies reporting Kaplan-Meier curves for cardiovascular risk or all-cause mortality. Individual patient data were reconstructed from published curves. Time-varying hazard ratios (HRs) were estimated using a mixed-effects Cox model with treatment-by-time interaction across pre-specified intervals (0–12, 12–24, 24–48, >48 months).

**Results:** Four studies (n=19,090) were included. Analysis revealed significant time-varying effects. For cardiovascular risk, HRs were 0.89 (95% CI 0.79 to 0.99) at 0–12 months, 0.58 (0.50 to 0.68) at 12–24 months, 0.86 (0.74 to 0.99) at 24–48 months, and 1.09 (0.81 to 1.46) after >48 months. For all-cause mortality, HRs were 0.70 (0.62 to 0.79), 0.39 (0.33 to 0.45), 0.75 (0.65 to 0.86), and 1.02 (0.77 to 1.34) across the same intervals, respectively.

**Conclusion:** The cardiovascular and mortality risk of febuxostat relative to allopurinol is time-dependent, showing a significant early reduction that attenuates over time. These findings advocate for a time-aware approach to clinical management and monitoring.

## 1. INTRODUCTION

Gout, a chronic inflammatory arthritis with rising global prevalence, poses a substantial burden on healthcare systems.^1, 2^ Beyond its debilitating articular manifestations, gout is a well-established independent risk factor for cardiovascular disease, making the cardiovascular safety of long-term urate-lowering therapy a critical concern in patient management.^3, 4^ Allopurinol has been the cornerstone of treatment, with febuxostat serving as an important alternative, particularly for allopurinol-intolerant patients.^5^

However, the cardiovascular safety of febuxostat has been a subject of intense debate and regulatory scrutiny following the US FDA’s black-box warning and the publication of the CARES trial, which suggested an increased risk of cardiovascular mortality compared to allopurinol.^6, 7^ In stark contrast, the larger and more pragmatic FAST trial found the two drugs to have comparable cardiovascular safety profiles.^8^ This discordance has created clinical uncertainty, reflected in cautious, conditional recommendations in major guidelines, such as the 2020 American College of Rheumatology guideline, which suggests considering a switch from febuxostat for patients with cardiovascular disease.^9^ Numerous observational studies and traditional meta-analyses have attempted to reconcile these findings with inconclusive and heterogeneous results.^10-14^ However, two major interrelated limitations in the existing literature preclude definitive conclusions. First, there has been a systematic neglect of temporal heterogeneity; the assumption of proportional hazards (a constant risk ratio over time) remains untested despite its profound clinical implications. Second, there is an artificial segregation of evidence types, failing to optimally integrate data from randomized controlled trials (RCTs) and high-quality real-world studies that use propensity score matching to control for confounding.^15^

Therefore, we conducted a time-to-event meta-analysis employing advanced individual patient data (IPD) reconstruction techniques. By integrating a comprehensive evidence base that includes both randomized trials and propensity score-matched observational cohorts, this study was explicitly designed to characterize and quantify the time-varying nature of the cardiovascular risk associated with febuxostat relative to allopurinol.

## 2. METHODS

This systematic review and meta-analysis was designed, conducted, and reported in adherence to the updated Preferred Reporting Items for Systematic Reviews and Meta-Analyses (PRISMA) 2020 statement.^16^ The study protocol received a priori prospective registration on the PROSPERO international register (registration identifier: CRD420251274101).

### 2.1 Eligible Criteria

Studies were selected based on the following pre-specified criteria. Participants: Adults with a definitive diagnosis of gout. Intervention and Comparator: Febuxostat (any dose) compared with allopurinol (any dose). Outcomes: The primary outcomes were time-to-event data for cardiovascular risk (defines as composite of cardiovascular death, nonfatal myocardial infarction, nonfatal stroke, urgent revascularization due to unstable angina, or hospitalization due to coronary heart failure) and all-cause mortality. Crucially, studies were required to present Kaplan-Meier (KM) curves derived from an intention-to-treat (ITT) analysis for these endpoints to be eligible for inclusion. Study Design: To comprehensively evaluate the cardiovascular risk profile of febuxostat, this analysis synthesizes evidence from both RCTs and high-quality observational studies employing propensity score matching (PSM).^15^ This approach acknowledges that while RCTs establish internal validity, well-conducted PSM studies improve external validity and complement RCT findings, enabling a more robust and generalizable safety assessment through meta-analysis.

### 2.2 Data Sources and Search Methods

A systematic and exhaustive literature search was executed to identify all relevant randomized controlled trials and prospective observational studies. Primary searches were performed in PubMed, Embase, and the Cochrane Library from their inception until November 4, 2025. The search strategy utilized a blend of subject headings (e.g., MeSH, Emtree) and free-text terms to capture studies pertaining to ‘gout,’ ‘febuxostat,’ and a comprehensive array of cardiovascular endpoints. To minimize publication bias and locate unpublished data, ClinicalTrials.gov was additionally scrutinized. The full electronic search strategies for all databases are available in the supplementary appendix. Manual screening of reference lists from eligible articles and relevant reviews supplemented the electronic search.

### 2.3 Selection Process, Data Extraction, and Individual Patient Data Reconstruction

Two reviewers independently screened records by title/abstract and subsequently by full text. Data extraction was independently performed by two researchers using a standardized electronic form. The extracted data encompassed three domains: (1) study-level descriptors (author, year, design, sample size), (2) aggregated patient baseline characteristics, and (3) outcome-specific time-to-event data; with any conflicts being resolved through consultation with a third, senior author.

For studies providing Kaplan-Meier (KM) curves, we implemented a robust, two-stage individual patient data (IPD) reconstruction methodology. First, digital curve images were systematically digitized using WebPlotDigitizer to obtain precise survival probabilities over time for each treatment arm. Second, these coordinates were algorithmically transformed into time-to-event datasets using the Guyot method within the IPDfromKM package.^17^ The details of IPD reconstruction provided in the Supplementary Materials.

### 2.4 Risk of Bias

The risk of bias assessment was performed independently by two investigators using standardized tools specific to each study design. For randomized controlled trials (RCTs), we employed the Cochrane Risk of Bias 2 (ROB-2) tool.^18^ For the included non-randomized studies (i.e., propensity score-matched observational studies), we utilized the Risk Of Bias In Non-randomized Studies – of Interventions (ROBINS-I), Version 2.^19^ Any discrepancies in the assessments between the two reviewers were resolved through discussion and, if necessary, by adjudication from a third senior methodologist to reach a consensus.

### 2.5 Statistical Analysis

The cumulative incidence of heart failure over time was initially described using the reconstructed Kaplan-Meier (KM) curves. The primary analysis was a one-stage individual patient data (IPD) meta-analysis using a mixed-effects Cox proportional hazards model, incorporating a random study intercept to account for between-study heterogeneity in baseline hazards.^20, 21^ To evaluate time-varying treatment effects, the model included an interaction term between treatment (febuxostat vs. allopurinol) and predefined follow-up intervals (to analyze time-varying treatment effects, follow-up time was stratified into four intervals based on the maximum follow-up across included studies: 0–12 months, 12–24 months, 24–48 months, and >48 months.). The significance of this interaction was tested via a likelihood ratio test. Hazard ratios (HRs) with 95% confidence intervals (CIs) were estimated for each interval, and corresponding absolute risk differences per 1000 patient-years were calculated.

Heterogeneity was assessed at three levels: baseline risk variation, consistency of the treatment effect across studies, and its stability within each time interval. A subgroup analysis compared estimates from RCTs and observational studies.

The robustness of the findings was evaluated through multiple sensitivity analyses: iterative exclusion of individual studies, a Bayesian survival model with MCMC sampling,^22^ and a traditional two-stage meta-analysis for methodological concordance.

### 2.6 Publication Bias

We assessed for publication bias and the effect of follow-up duration via meta-regression. Using study-level data from the two-stage analysis, we regressed the log hazard ratios against each study’s median follow-up time in a random-effects model (REML estimation).^23^

### 2.7 Certainty Assessment

To transparently grade the confidence in our findings, we applied the GRADE (Grading of Recommendations Assessment, Development, and Evaluation) framework.^24, 25^ This involved a sequential judgment of five pre-defined domains that may lower or raise the certainty of evidence: 1) risk of bias (study limitations), 2) inconsistency (heterogeneity), 3) indirectness, 4) imprecision, and 5) publication bias. An initial high-certainty rating for evidence from randomized trials was subsequently adjusted based on assessments within these domains, yielding a final grade for each outcome.

## 3. RESULTS

### 3.1 Study Selection and Baseline Characteristics

Our systematic search identified 348 records. After removing duplicates and screening titles/abstracts, 33 full-text articles were assessed. Eight studies were evaluated in detail, of which four were excluded for lacking necessary Kaplan-Meier curves (n=2) or ITT analysis (n=2). Consequently, four studies fulfilled all criteria and were included in the analysis (Figure 1).

**Figure 1.**
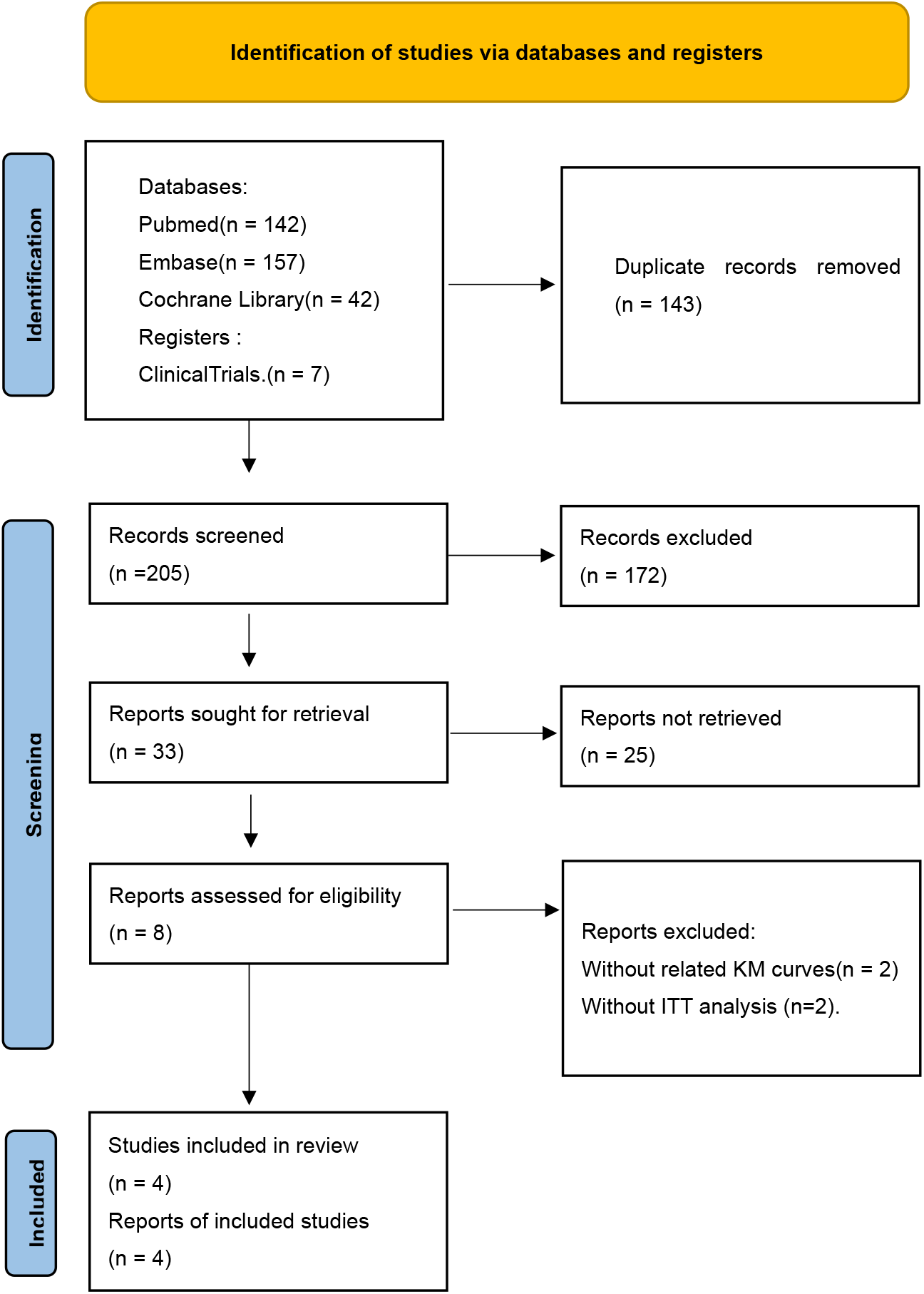
PRISMA Flow Diagram of Study Selection. A PRISMA flowchart summarizing the identification, screening, eligibility assessment, and inclusion of studies in the meta-analysis.

Table 1 details the baseline characteristics. Two RCTs (CARES, FAST) and two NRS (Ju C 2020, Yang 2022) were included.^7, 8, 26, 27^ The RCTs had larger sample sizes than the NRS. Patient age was consistently high (median/mean 64-71 years). The male percentage ranged from 58.7% to 85.5%. Follow-up duration varied from a median of 19.3 months to 1467 days.

### 3.2 Risk of Bias Assessment

Risk of bias assessments are detailed in Supplementary Section 3. For RCTs (ROB-2), the FAST trial had a low overall risk, while the CARES trial had a high overall risk, mainly due to missing data. For non-randomized studies (ROBINS-I V2), Ju C et al. had a moderate overall risk, and Yang et al. had a serious overall risk due to confounding and selection issues.

### 3.3 Results of Reconstructed Kaplan-Meier curves

The reconstructed Kaplan-Meier curves for the composite cardiovascular endpoint (MACE), integrating individual patient data from all four studies, are presented in Figure 2A. Visual inspection suggests an early separation of the curves. Contrary to the initial impression from some individual trials, the pooled analysis indicates that the cumulative incidence in the febuxostat group appears comparable to or marginally higher than that in the allopurinol group during the initial follow-up period. The two curves subsequently converge over longer follow-up. The log-rank test comparing the overall survival distributions across the entire follow-up period yielded a p-value of 0.074, which did not reach conventional statistical significance.

**Figure 2.**
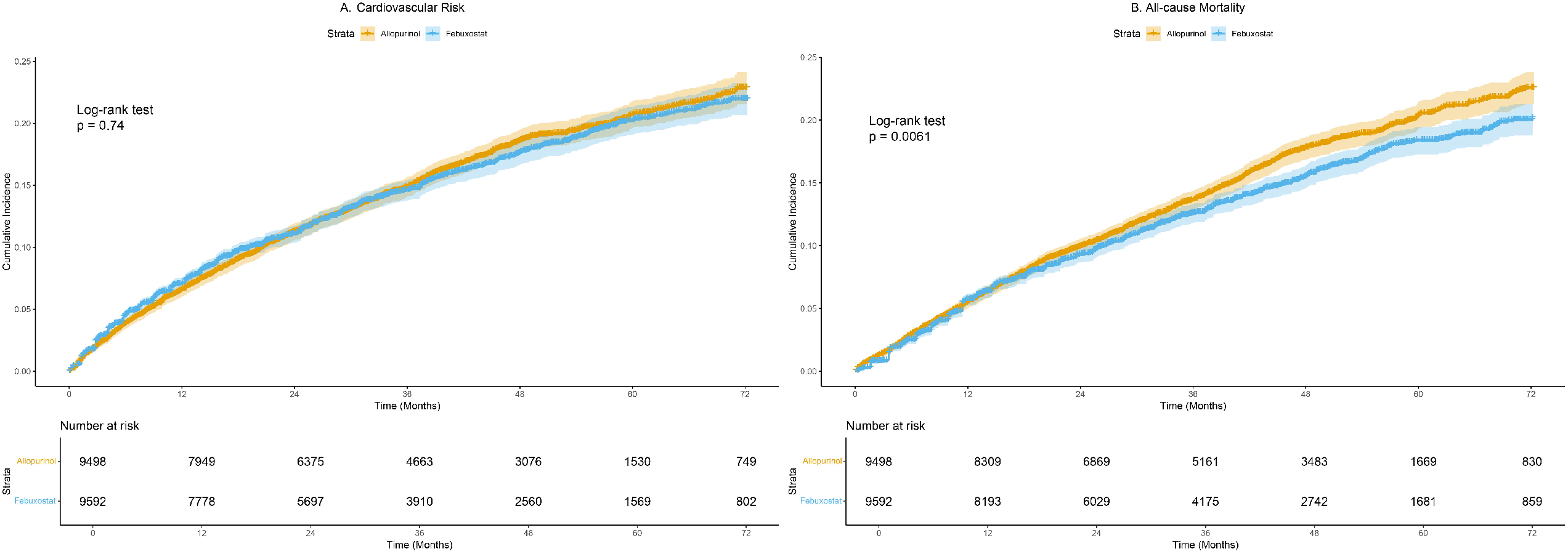
Reconstructed Kaplan-Meier Curves for Primary Outcomes. Pooled time-to-event curves for the (A) composite cardiovascular endpoint and (B) all-cause mortality, derived from reconstructed individual patient data from all four included studies. The solid line represents the febuxostat group, and the dashed line represents the allopurinol group. Shaded areas denote the 95% confidence intervals. The p-values are from the log-rank test comparing the overall survival distributions between treatment groups. The number of patients at risk at specified time points is indicated below each panel.

For all-cause mortality, the pooled Kaplan-Meier curves demonstrate a clear and early separation (Figure 2B). The curve for the febuxostat group shows a lower cumulative incidence of death compared to the allopurinol group from the outset, and this difference appears to widen over time. The log-rank test substantiated this visual interpretation with a statistically significant p-value of 0.004. Study-specific reconstructed curves are available in Supplementary Materials, Section 8.

### 3.4 Results of Main Analysis

Our primary time-varying analysis demonstrated a significant, non-linear relationship between treatment and cardiovascular risk (Figure 3A). Febuxostat was associated with a lower hazard in the early-to-mid follow-up periods: HR 0.89 (95% CI 0.79–0.99) for 0–12 months, HR 0.58 (95% CI 0.50–0.68) for 12–24 months, and HR 0.86 (95% CI 0.74–0.99) for 24–48 months. Beyond 48 months, the direction of effect reversed, showing a non-significant increased risk (HR 1.09, 95% CI 0.81–1.46). The corresponding absolute risk reductions per 1000 patient-years were 8, 22, and 7 fewer events in the first three intervals, respectively. The certainty of evidence was graded as Moderate for the 0–12 month interval and Low for subsequent intervals, primarily due to risk of bias and inconsistency (Table 2).

**Figure 3.**
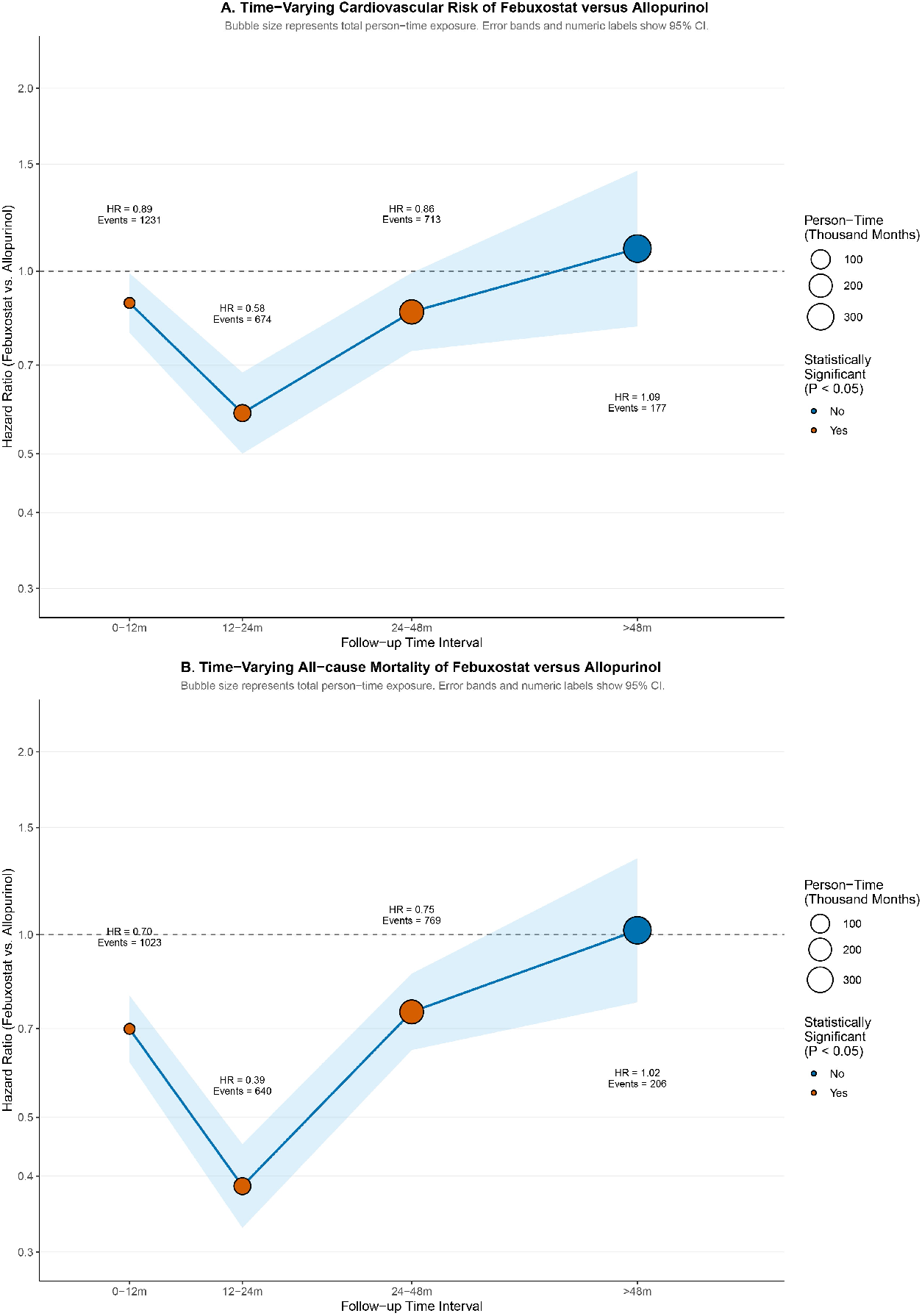
Interval-Specific Hazard Ratios from the Primary Mixed-Effects Cox Model. Graphical output of the one-stage IPD meta-analysis modeling a treatment-by-time interaction. Estimates are presented for four consecutive intervals post-treatment initiation. For each interval in Panels A (Composite cardiovascular endpoint) and B (Mortality), the hazard ratio (point estimate), 95% CI, total event count, and proportional person-time exposure (bubble area) are displayed. The model reveals significant departure from proportional hazards.

For all-cause mortality, the time-varying hazard ratios exhibited a similar but more pronounced pattern (Figure 3B). Febuxostat use was linked to a substantially lower risk of death in the first four years, with a near 60% reduction at 12–24 months: HR 0.70 (95% CI 0.62–0.79) for 0–12 months, HR 0.39 (95% CI 0.33–0.45) for 12–24 months, and HR 0.75 (95% CI 0.65–0.86) for 24–48 months. In the >48 month interval, no significant difference was detected (HR 1.02, 95% CI 0.77–1.34). The absolute risk differences corresponded to 17, 29, and 11 fewer deaths per 1000 patient-years in the respective intervals showing benefit. The certainty of evidence for all mortality outcomes was rated as Low, downgraded for risk of bias and inconsistency (Table 2).

### 3.5 Validation of Synthesis Robustness

Assessment of heterogeneity revealed statistical inconsistency in the pooled estimates. For baseline risk, the I^2^ statistics were 9.0% for cardiovascular outcomes and 14.7% for all-cause mortality. For treatment effect heterogeneity, the I^2^ values were 1.0% and 7.3%, respectively. Time-stratified and subgroup analyses by study type suggested that the observed minor heterogeneity was likely statistical in nature and largely attributable to the methodological profile of the CARES trial, as detailed in Supplementary Materials, Section 5.

The robustness of the primary findings was confirmed through three sensitivity analyses. All approaches yielded effect estimates consistent in direction and magnitude with the primary one-stage IPD model, affirming the stability of our conclusions (Supplementary Materials, Section 6). Furthermore, the lack of detectable publication bias strengthens the credibility of the synthesized evidence (Supplementary Materials, Section 7).

## 4. DISCUSSION

This one-stage IPD meta-analysis, synthesizing patient-level data from over 19,000 individuals, demonstrates a fundamental and previously unquantified aspect: the cardiovascular and mortality risk profile of febuxostat relative to allopurinol is intrinsically dynamic, not static. Our model reveals a significant temporal heterogeneity, with febuxostat associated with a substantially lower hazard during the initial 48 months of treatment—most markedly between 1-2 years—followed by a period of statistical equipoise beyond four years. These findings move beyond the binary debate of ‘safe versus unsafe’ and provides a coherent, time-stratified framework to reinterpret the entire evidence landscape.

The stark dichotomy between the CARES and FAST trials has been a source of clinical paralysis. Our analysis suggests this conflict may be partially artifactual, arising from comparing aggregate outcomes over different segments of a non-proportional hazard curve. CARES, with its concerning signal, may have been disproportionately influenced by late-occurring events in a trial plagued by exceptionally high attrition (>45%), which critically compromises the interpretability of its time-to-event analyses.^7^ FAST, a more pragmatic trial with robust follow-up, may have better captured the earlier period where benefit predominates. It is crucial to note that the primary alarm from CARES was specific to cardiovascular mortality.^8^ In our exploratory analysis of this endpoint (Supplementary Section 9), which was only feasible with the two RCTs, hazard ratios across all predefined intervals were non-significant. However, a telling pattern emerged: the point estimates progressively increased from below 1.0 in the early intervals to above 1.0 in the >48-month window. While statistically inconclusive, this trend underscores that the early composite benefit we observed is not necessarily driven by a reduction in cardiovascular death, and that the long-term signal warrants vigilant monitoring. It also highlights why the CARES findings, while important, should not be the sole anchor for guidelines, given their methodological vulnerability.

Our study represents a deliberate advance beyond traditional meta-analyses, which have consistently produced conflicting single-point estimates. By forcing a constant hazard ratio, these syntheses obscured the true nature of the risk relationship. The work of Guan et al. was a seminal step for employing IPD reconstruction; however, by limiting inclusion to two RCTs and reporting only a cumulative, non-significant effect, it could not unveil the temporal pattern.^28^ Our integration of propensity-score matched observational studies added crucial statistical power and clinical diversity, enabling our pre-specified, interval-based model to detect and quantify the significant early advantage. This is not a minor methodological tweak but a paradigm shift in evidence synthesis for drug safety, moving from asking “*What is the average risk*?” to “*How does risk change over time*?”.

The finding of an early cardiovascular benefit for febuxostat is mechanistically plausible. Febuxostat provides a rapid, potent, and dose-independent reduction in serum urate.^29^ This swift decline may lead to a quicker suppression of the chronic, low-grade inflammation and endothelial dysfunction mediated by soluble urate and crystalline deposits, potentially translating into an accelerated reduction in cardiovascular stress compared to the slower titration often required with allopurinol.^30^ This has direct and challenging implications for clinical guidelines. The 2020 ACR guideline, influenced heavily by CARES, conditionally recommends considering a switch *from* febuxostat *to* an alternative in patients with cardiovascular disease.^9^ Our analysis, which included populations with significant cardiovascular comorbidity (indeed, CARES enrolled only such patients), directly contests the universality of this advice. For a patient initiating therapy, especially one at high short-term cardiovascular risk, our data suggest febuxostat could be a rational and potentially protective choice. The clinical imperative should therefore shift from a blanket switching recommendation to a structured, time-strategic management plan: initiating febuxostat with confidence in its early profile, while implementing scheduled, comprehensive cardiovascular risk reassessments as treatment extends beyond 3-4 years.

Our study has several limitations. First, while validated, IPD reconstruction from curves is an approximation. Second, despite propensity score matching, unmeasured confounding in the observational studies cannot be ruled out. Third, and most significant for clinical application, the patient numbers and event counts in the >48-month interval were sparse, rendering our estimates for long-term use imprecise and necessitating caution. The analysis also remains partially anchored to the high-risk-of-bias CARES trial.

Despite these limitations, the study’s strengths are substantial and reinforce the primary conclusion. The use of a one-stage mixed-effects Cox model formally accounted for between-study heterogeneity. The pre-specified testing of the time-treatment interaction provides robust statistical evidence against proportional hazards. The methodological triangulation—through leave-one-out, Bayesian, and two-stage sensitivity analyses—all converged on the same temporal pattern, indicating the finding is not an artifact of a single analytical approach. Furthermore, by integrating RCTs with real-world evidence, we enhanced the external validity and power of our time-stratified estimates.^31^

## 5. CONCLUSION

In summary, febuxostat’s cardiovascular risk profile relative to allopurinol is time-dependent. The evidence suggests a favorable window in the early-to-mid phase of therapy, shifting to a less certain profile with prolonged use. Therefore, a nuanced, time-strategic approach to management is recommended, moving away from binary classifications of safety.

## DATA AVAILABILITY STATEMENT

The original contributions presented in the study are included in the article/Supplementary Materials, further inquiries can be directed to the corresponding author.

## HUMAN ETHICS AND CONSENT TO PARTICIPATE DECLARATIONS

Not applicable. This meta-analysis utilized only data extracted from previously published studies via Kaplan-Meier curve reconstruction. It did not involve any direct access to individual patient records or identifiable data.

## AUTHOR CONTRIBUTIONS

QW, YS, RH, HW, YL, JZ and SZ were in charge of study design, QW, YS, and SZ completed data collection and interpretation, assessment of certainty. QW, YS, RH wrote this manuscript together, all authors critically reviewed the manuscript and provided revision, SZ was the primary monitor of this study. YL, LL, DS, and JZ mainly checked the final data of the study. All authors contributed to the article and approved the submitting version.

