## Supplementary material for "Time-Varying Cardiovascular Risk of Febuxostat versus Allopurinol in Gout: A One-stage Meta-analysis": Table 1

**Table 2. Baseline Characteristics of Included Studies**

| **Study ID** | **Design** | **No. of Patients** | | **Age** | | **Male (%)** | | **Follow-up Duration** |
| --- | --- | --- | --- | --- | --- | --- | --- | --- |
|  |  | **Febuxostat** | **Allopurinol** | **Febuxostat** | **Allopurinol** | **Febuxostat** | **Allopurinol** |  |
| **CARES** | RCT | 3098 | 3092 | 64 (58–71) | 65 (58–71) | 84.10% | 83.80% | 32 months |
| **FAST** | RCT | 3063 | 3065 | 71.0±6.4 | 70.9±6.5 | 85.50% | 85.00% | 1467 days |
| **Ju C 2020** | NRS | 276 | 828 | 70.41(14.35) | 70.01(14.90) | 67.40% | 66.30% | 1.97 years |
| **Yang 2022** | NRS | 3155 | 2513 | 70 (12) | 70 (12) | 58.72% | 59.12% | 19 months |
