## Supplementary material for "Time-Varying Cardiovascular Risk of Febuxostat versus Allopurinol in Gout: A One-stage Meta-analysis": Table 2

**Table 2. GRADE Summary of Finding Tables**

| **Outcomes** | **Time Intervals** | **Febuxostat vs Allopurinol Hazard Ratio  (95% Confidence Interval)** | **Anticipated absolute effect  (95% confidence interval) per 1000 patient-years** | | **Risk Difference (95% Confidence Interval) per 1000 patient-years** | **Certainty of Evidence** |
| --- | --- | --- | --- | --- | --- | --- |
|  |  |  | **With Allopurinol** | **With Febuxostat** |  |  |
| **Cardiovascular Endpoints** | 0-12 months | 0.89 (0.79, 0.99) | 68 | 60 (54 to 67) | 8 fewer (14 fewer to 1 fewer) | Moderate ^1^ |
|  | 12-24 months | 0.58 (0.50, 0.68) | 52 | 30 (26 to 36) | 22 fewer (26 fewer to 16 fewer) | Low ^1, 2^ |
|  | 24-48 months | 0.86 (0.74, 0.99) | 43 | 36 (32 to 42) | 7 fewer (11 fewer to 1 fewer) | Low ^1, 2^ |
|  | > 48 months | 1.09 (0.81, 1.46) | 26 | 29 (21 to 39) | 3 more (5 fewer to 13 more) | Low ^1, 3^ |
| **All-cause mortality** | 0-12 months | 0.70 (0.62, 0.79) | 57 | 40 (35 to 45) | 17 fewer (22 fewer to 12 fewer) | Low ^1, 2^ |
|  | 12-24 months | 0.39 (0.33, 0.45) | 48 | 19 (16 to 22) | 29 fewer (32 fewer to 26 fewer) | Low ^1, 2^ |
|  | 24-48 months | 0.75 (0.65, 0.86） | 45 | 34 (29 to 39) | 11 fewer (16 fewer to 6 fewer) | Low ^1, 2^ |
|  | > 48 months | 1.02 (0.77, 1.34) | 30 | 30 (23 to 40) | 0 more (7 fewer to 10 more) | Low ^1, 3^ |

Notes: Reasons for rating down: 1. risk of bias; 2. inconsistency; 3.imprecision
