## Supplemental Materials for "Time-Varying Cardiovascular Risk of Febuxostat versus Allopurinol in Gout: A One-stage Meta-analysis"

### Supplementary Materials: Time Dependent Cardiovascular Risk of Febuxostat versus Allopurinol in Gout: A One-stage Meta-analysis

#### 1. PRISMA Checklist

| Section and Topic | Item # | Checklist item | Location where item is reported |
|---|---|---|---|
| **TITLE** | | | |
| Title | 1 | Identify the report as a systematic review. | Lines 1-2 |
| **ABSTRACT** | | | |
| Abstract | 2 | See the PRISMA 2020 for Abstracts checklist. | Lines 21-45 |
| **INTRODUCTION** | | | |
| Rationale | 3 | Describe the rationale for the review in the context of existing knowledge. | Lines 50-70 |
| Objectives | 4 | Provide an explicit statement of the objective(s) or question(s) the review addresses. | Lines 71-75 |
| **METHODS** | | | |
| Eligibility criteria | 5 | Specify the inclusion and exclusion criteria for the review and how studies were grouped for the syntheses. | Section 2.1 |
| Information sources | 6 | Specify all databases, registers, websites, organisations, reference lists and other sources searched or consulted to identify studies. Specify the date when each source was last searched or consulted. | Section 2.2 |
| Search strategy | 7 | Present the full search strategies for all databases, registers and websites, including any filters and limits used. | Section 2.2 and supplementary materials section 2 |
| Selection process | 8 | Specify the methods used to decide whether a study met the inclusion criteria of the review, including how many reviewers screened each record and each report retrieved, whether they worked independently, and if applicable, details of automation tools used in the process. | Section 2.3 |
| Data collection process | 9 | Specify the methods used to collect data from reports, including how many reviewers collected data from each report, whether they worked independently, any processes for obtaining or confirming data from study investigators, and if applicable, details of automation tools used in the process. | Section 2.3 |

| Section and Topic | Item # | Checklist item | Location where item is reported |
|---|---|---|---|
| Data items | 10a | List and define all outcomes for which data were sought. Specify whether all results that were compatible with each outcome domain in each study were sought (e.g. for all measures, time points, analyses), and if not, the methods used to decide which results to collect. | Section 2.3 |
| | 10b | List and define all other variables for which data were sought (e.g. participant and intervention characteristics, funding sources). Describe any assumptions made about any missing or unclear information. | Section 2.3 |
| Study risk of bias assessment | 11 | Specify the methods used to assess risk of bias in the included studies, including details of the tool(s) used, how many reviewers assessed each study and whether they worked independently, and if applicable, details of automation tools used in the process. | Section 2.4 |
| Effect measures | 12 | Specify for each outcome the effect measure(s) (e.g. risk ratio, mean difference) used in the synthesis or presentation of results. | Section 2.5 |
| Synthesis methods | 13a | Describe the processes used to decide which studies were eligible for each synthesis (e.g. tabulating the study intervention characteristics and comparing against the planned groups for each synthesis (item #5)). | Section 2.5 |
| | 13b | Describe any methods required to prepare the data for presentation or synthesis, such as handling of missing summary statistics, or data conversions. | Section 2.5 |
| | 13c | Describe any methods used to tabulate or visually display results of individual studies and syntheses. | Section 2.5 |
| | 13d | Describe any methods used to synthesize results and provide a rationale for the choice(s). If meta-analysis was performed, describe the model(s), method(s) to identify the presence and extent of statistical heterogeneity, and software package(s) used. | Section 2.5 |
| | 13e | Describe any methods used to explore possible causes of heterogeneity among study results (e.g. subgroup analysis, meta-regression). | Section 2.5 |
| | 13f | Describe any sensitivity analyses conducted to assess robustness of the synthesized results. | Section 2.5 |
| Reporting bias assessment | 14 | Describe any methods used to assess risk of bias due to missing results in a synthesis (arising from reporting biases). | Section 2.6 |
| Certainty assessment | 15 | Describe any methods used to assess certainty (or confidence) in the body of evidence for an outcome. | Section 2.7 |
| **RESULTS** | | | |

| Section and Topic | Item # | Checklist item | Location where item is reported |
|---|---|---|---|
| Study selection | 16a | Describe the results of the search and selection process, from the number of records identified in the search to the number of studies included in the review, ideally using a flow diagram. | Section 3.1 |
| | 16b | Cite studies that might appear to meet the inclusion criteria, but which were excluded, and explain why they were excluded. | Section 3.1 |
| Study characteristics | 17 | Cite each included study and present its characteristics. | Table 1 |
| Risk of bias in studies | 18 | Present assessments of risk of bias for each included study. | Section 3.2 |
| Results of individual studies | 19 | For all outcomes, present, for each study: (a) summary statistics for each group (where appropriate) and (b) an effect estimate and its precision (e.g. confidence/credible interval), ideally using structured tables or plots. | Supplementary materials section 6 and 8 |
| Results of syntheses | 20a | For each synthesis, briefly summarise the characteristics and risk of bias among contributing studies. | Section 3.4 |
| | 20b | Present results of all statistical syntheses conducted. If meta-analysis was done, present for each the summary estimate and its precision (e.g. confidence/credible interval) and measures of statistical heterogeneity. If comparing groups, describe the direction of the effect. | Section 3.4 |
| | 20c | Present results of all investigations of possible causes of heterogeneity among study results. | Lines 197-202 and supplementary materials section 5 |
| | 20d | Present results of all sensitivity analyses conducted to assess the robustness of the synthesized results. | Lines 203-205 and supplementary materials section 6 |
| Reporting biases | 21 | Present assessments of risk of bias due to missing results (arising from reporting biases) for each synthesis assessed. | Lines 206-207 and supplementary materials section 7 |

| Section and Topic | Item # | Checklist item | Location where item is reported |
|---|---|---|---|
| Certainty of evidence | 22 | Present assessments of certainty (or confidence) in the body of evidence for each outcome assessed. | Table 2 |
| **DISCUSSION** | | | |
| Discussion | 23a | Provide a general interpretation of the results in the context of other evidence. | Lines 209-212 |
| | 23b | Discuss any limitations of the evidence included in the review. | Lines 226-228 |
| | 23c | Discuss any limitations of the review processes used. | Lines 228-233 |
| | 23d | Discuss implications of the results for practice, policy, and future research. | Lines 213-225 |
| **OTHER INFORMATION** | | | |
| Registration and protocol | 24a | Provide registration information for the review, including register name and registration number, or state that the review was not registered. | Lines 77-80 |
| | 24b | Indicate where the review protocol can be accessed, or state that a protocol was not prepared. | |
| | 24c | Describe and explain any amendments to information provided at registration or in the protocol. | |
| Support | 25 | Describe sources of financial or non-financial support for the review, and the role of the funders or sponsors in the review. | Lines 264-268 |
| Competing interests | 26 | Declare any competing interests of review authors. | Lines 261-263 |
| Availability of data, code and other materials | 27 | Report which of the following are publicly available and where they can be found: template data collection forms; data extracted from included studies; data used for all analyses; analytic code; any other materials used in the review. | Lines 248-250 |

#### 2. Search Strategies

PubMed:

#1 “Gout”[Mesh] OR gout

#2 “febuxostat”[Mesh] OR febuxostat

#3 stroke OR "myocardial infarction" OR mortality OR "cardiovascular mortality" OR "cardiovascular risk" OR "cardiovascular safety" OR cardiovascular OR cardio OR heart OR "Cardiology"[Mesh]

#4 "Observational Study"[Publication Type] AND "Observational Studies as Topic"[Mesh]

#5 observational OR cohort OR "Cohort Studies"[Mesh]

#6 "Randomized Controlled Trials as Topic"[Mesh] OR "Randomized Controlled Trial"[Publication Type]

#7 #4 OR #5 OR #6

#8 #1 AND #2 AND #3 AND #7

Embase:

#1 'gout'/exp OR 'gout'

#2 'febuxostat'/exp OR 'febuxostat'

#3 'cardiovascular disease'

#4 'major adverse cardiovascular events'

#5 'mortality'/exp OR 'mortality'

#6 'heart infarction'/exp OR 'heart infarction'

#7 'cerebrovascular accident'/exp OR 'cerebrovascular accident'

#8 'randomized controlled trial'/exp OR 'randomized controlled trial'

#9 'observational study'/exp OR 'observational study'

#1 AND #2 AND (#3 OR #4 OR #5 OR #6 OR #7) AND (#8 OR #9)

Cochrane Library:

gout AND febuxostat AND ("major adverse cardiac event" OR mortality OR "myocardial infarction" OR stroke OR “death” OR “mortality”)

#### 3. Risk of Bias

##### 3.1 ROB 2.0 for Included RCTs

| Study ID | Randomization Process | Deviations from the intended interventions | Missing outcome data | Measurement of the outcome | Selection of the reported result | Overall |
| --- | --- | --- | --- | --- | --- | --- |
| CARES | Low risk | Some concerns | High risk | Low risk | Low risk | High risk |
| FAST | Low risk | Some concerns | Low risk | Low risk | Low risk | Low risk |

##### 3.2 ROBINS-I V2 for Included NRSs

| Study ID | Bias due to confounding | Bias in classification of interventions | Bias in selection of participants into the study | Bias due to missing data | Bias in measurement of the outcome | Bias in selection of the report results | Overall |
| --- | --- | --- | --- | --- | --- | --- | --- |
| Ju C 2020 | Low | Low | Low | Low | Low | Moderate | Moderate |
| Yang 2022 | Serious | Low | Serious | Low | Low | Moderate | Serious |

#### 4. Details of Methods

##### 4.1 Workflow

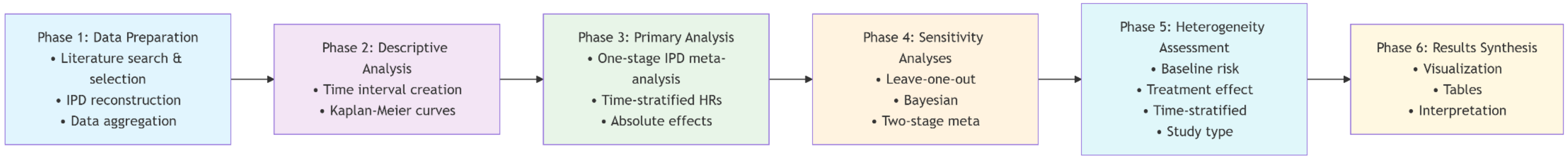

##### 4.2 Individual Patient Data (IPD)reconstruction

For studies that published Kaplan-Meier curves without providing individual-level data, we reconstructed the IPD using a two-step process. First, we extracted coordinate data from digital images of published Kaplan-Meier curves using WebPlotDigitizer version 4.7 (Automeris, USA). This semi-automated tool enabled precise extraction of time points and corresponding survival probabilities at regular intervals along the curves. For each study, we extracted data separately for the allopurinol (control) and febuxostat (intervention) groups, ensuring consistency in the time scale and survival probability measurements.

Subsequently, we employed the IPDfromKM R package (version 0.1.10) to reconstruct individual-level time-to-event data from the extracted coordinates. The algorithm within this package utilizes the Guyot transformation method, which reconstructs individual event times by simulating censoring distributions consistent with the reported number at risk and survival probabilities. The reconstruction process accounted for the number of patients at risk at various time points when such information was available in the original publications. For studies reporting propensity score-matched cohorts, we maintained the matched structure in the reconstructed data by preserving the reported sample sizes and event counts.

4.3 Primary analysis

The primary analysis was a one-step individual patient data meta-analysis using a mixed-effects Cox proportional hazards model, which accounts for between-study heterogeneity in baseline hazards. The model was specified with a random intercept for study to allow baseline hazards to vary across studies. To capture time-varying treatment effects, we included an interaction term between treatment and the predefined time intervals. The model was fitted using the coxme package in R.

Specifically, the model was defined as:

$$h_{ij}(t) = h_{0j}(t)\exp\left(\beta_1 \text{Treatment}_{ij} + \beta_2 \text{TimeInterval}_{ij} + \beta_3 \text{Treatment}_{ij} \times \text{TimeInterval}_{ij} + u_j\right)$$

where $h_{ij}(t)$ is the hazard for patient $i$ in study $j$, $h_{0j}(t)$ is the study-specific baseline hazard, $\text{Treatment}_{ij}$ is the treatment indicator (0: allopurinol, 1: febuxostat), $\text{TimeInterval}_{ij}$ is a categorical variable for the time intervals, and $u_j \sim N(0, \tau^2)$ is the random study effect.

We tested the significance of the time-varying effect by comparing the model with and without the treatment-by-time interaction using a likelihood ratio test. Hazard ratios (HRs) for febuxostat versus allopurinol were calculated for each time interval along with 95% confidence intervals (CIs).

Absolute effects were calculated as the number of events per 1000 patient-years. For each time interval, we computed the incidence rate in the allopurinol group (control) by dividing the number of events by the total person-years in that interval and multiplying by 1000. Person-years were calculated by summing the follow-up time (converted to years) contributed by all patients in the control group within each interval. The incidence rate in the febuxostat group was then estimated by multiplying the control incidence rate by the HR for that interval. The absolute risk difference (ARD) was calculated as the difference between the febuxostat and allopurinol incidence rates.

4.4 Heterogeneity assessment

We assessed heterogeneity in multiple dimensions:

Baseline risk heterogeneity: The variance of the random intercept ($\tau^2$) in the primary mixed-effects Cox model was used to quantify between-study heterogeneity in baseline hazards. The $I^2$ statistic was derived by comparing this variance to the typical within-study variance ($\pi^2/6$).

Treatment effect heterogeneity: We fitted an additional mixed-effects Cox model that included a random slope for treatment, allowing the treatment effect to vary across studies. The likelihood ratio test was used to compare this model with the primary model (random intercept only) to determine if there was significant variability in treatment effects.

Time-stratified heterogeneity: For each time interval, we performed a two-stage random-effects meta-analysis of study-specific HRs (estimated from Cox models within each study for that interval) to compute interval-specific $I^2$ statistics. This allowed us to assess how heterogeneity varied over time.

Subgroup analysis by study type: We compared the treatment effects between RCTs and NRS by including an interaction term between treatment and study type in the primary model. Additionally, we performed separate one-stage IPD meta-analyses for RCTs and NRS to obtain pooled HRs for each subgroup.

4.5 Sensitivity analyses

We performed several sensitivity analyses to assess the robustness of the primary analysis results.

Leave-one-out analysis: We repeated the primary analysis by excluding one study at a time to evaluate whether any single study disproportionately influenced the overall results.

Bayesian mixed-effects model: We fitted a Bayesian parametric survival model using the brms package. A Weibull distribution was assumed for the survival times, with weakly informative priors for the fixed effects (normal distribution with mean 0 and standard deviation 2.5) and random effects (student-t distribution with 3 degrees of freedom). The model included the same treatment-by-time interaction and random study intercept as the primary analysis. Markov chain Monte Carlo (MCMC) sampling was performed with three chains, each with 3000 iterations (1500 warm-up). Posterior distributions of HRs for each time interval were summarized with medians and 95% credible intervals.

Two-stage meta-analysis: For methodological comparison, we also conducted a conventional two-stage meta-analysis ignoring time-varying effects. In the first stage, we fitted a Cox proportional hazards model within each study to obtain study-specific log HRs and standard errors. In the second stage, these estimates were combined using a random-effects meta-analysis model (restricted maximum likelihood estimator) via the metafor package. Heterogeneity was quantified by the $I^2$ statistic. Additionally, a meta-regression was performed to explore whether the median follow-up time of each study explained heterogeneity in treatment effects.

#### 5. Results of Heterogeneity Assessment

##### 5.1 Heterogeneity assessment of cardiovascular endpoints

Baseline risk heterogeneity: $\tau^2$ = 0.164, $I^2$ = 9.0%.

Treatment effect heterogeneity: $\tau^2$ = 0.017, $I^2$ = 1.0%.

Figure S1. Heterogeneity across time intervals for cardiovascular endpoints

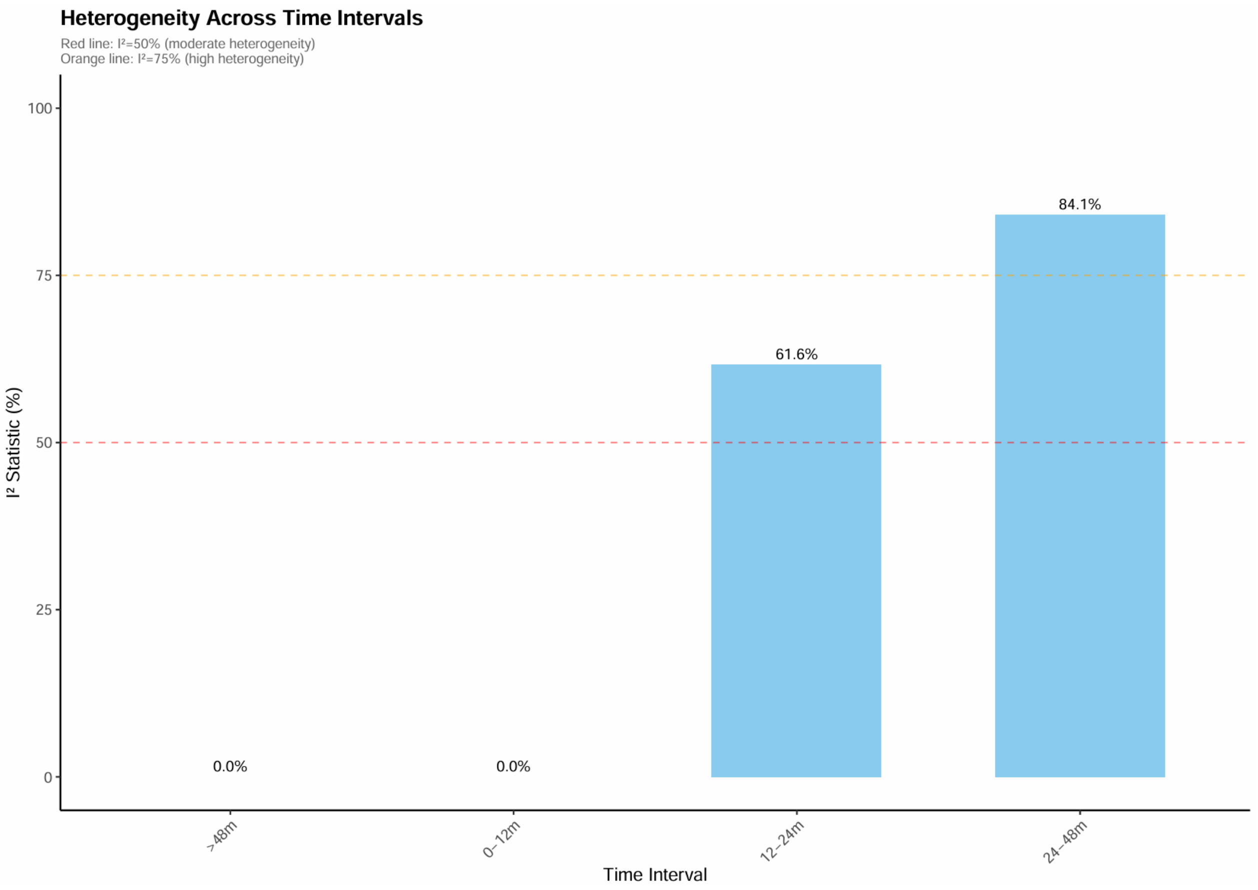

Figure S2. Subgroup analysis by study type for cardiovascular endpoints

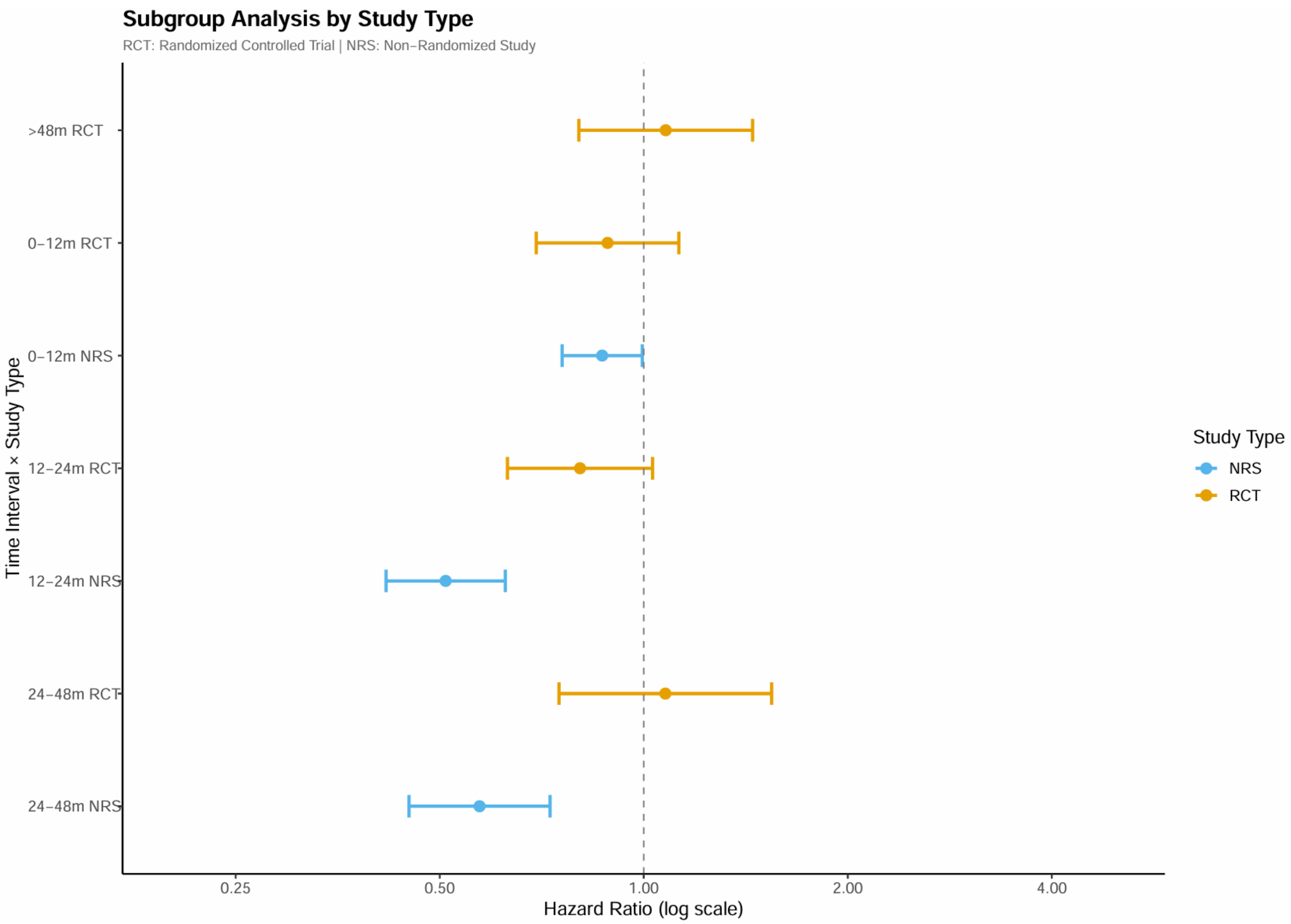

5.2 Heterogeneity assessment of all-cause mortality

Baseline risk heterogeneity: $\tau^2$ = 0.273, $I^2$ = 14.7%.

Treatment effect heterogeneity: $\tau^2$ = 0.129, $I^2$ = 7.3%.

Figure S3. Heterogeneity across time intervals for all-cause mortality

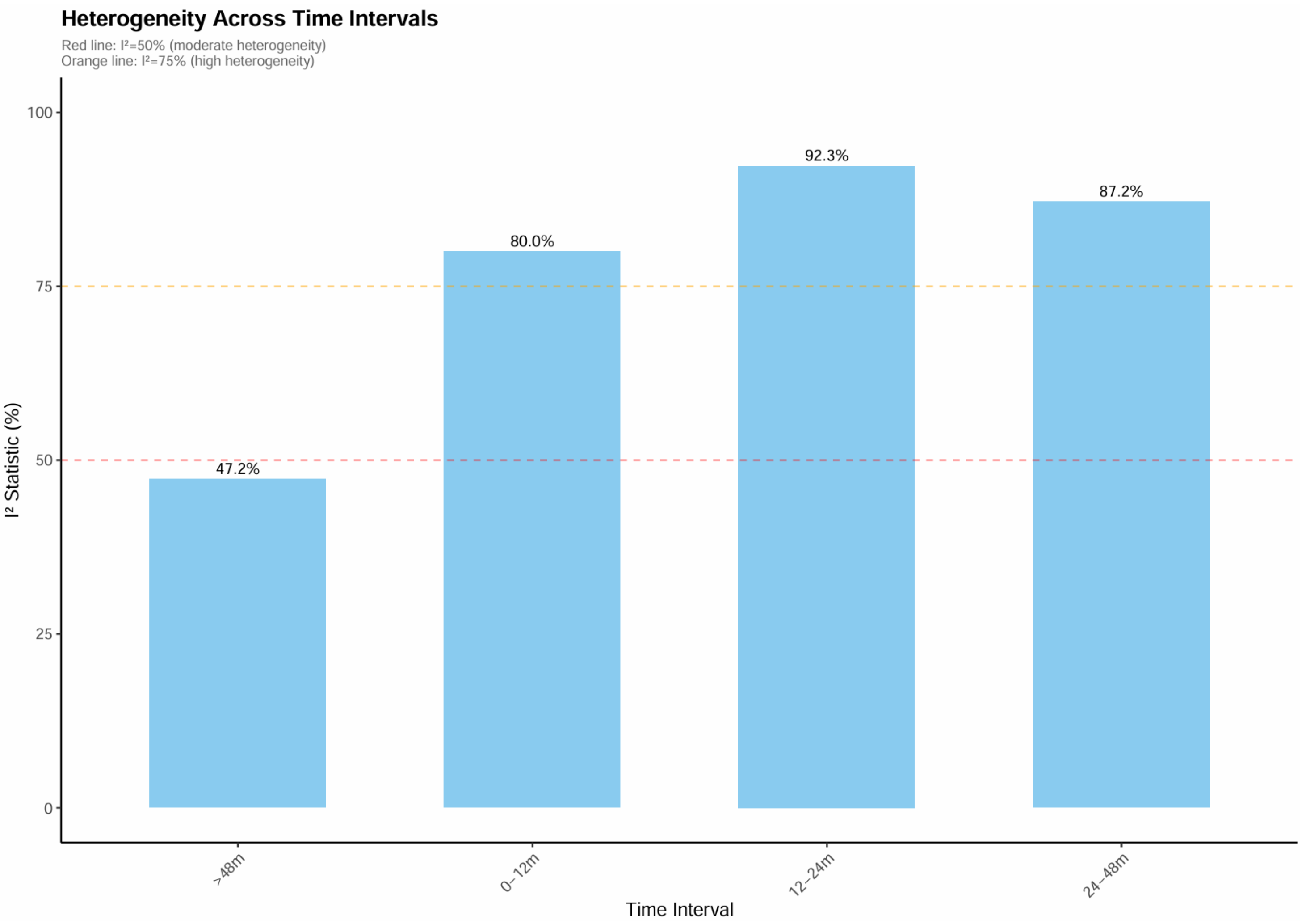

Figure S4. Subgroup analysis by study type for all-cause mortality

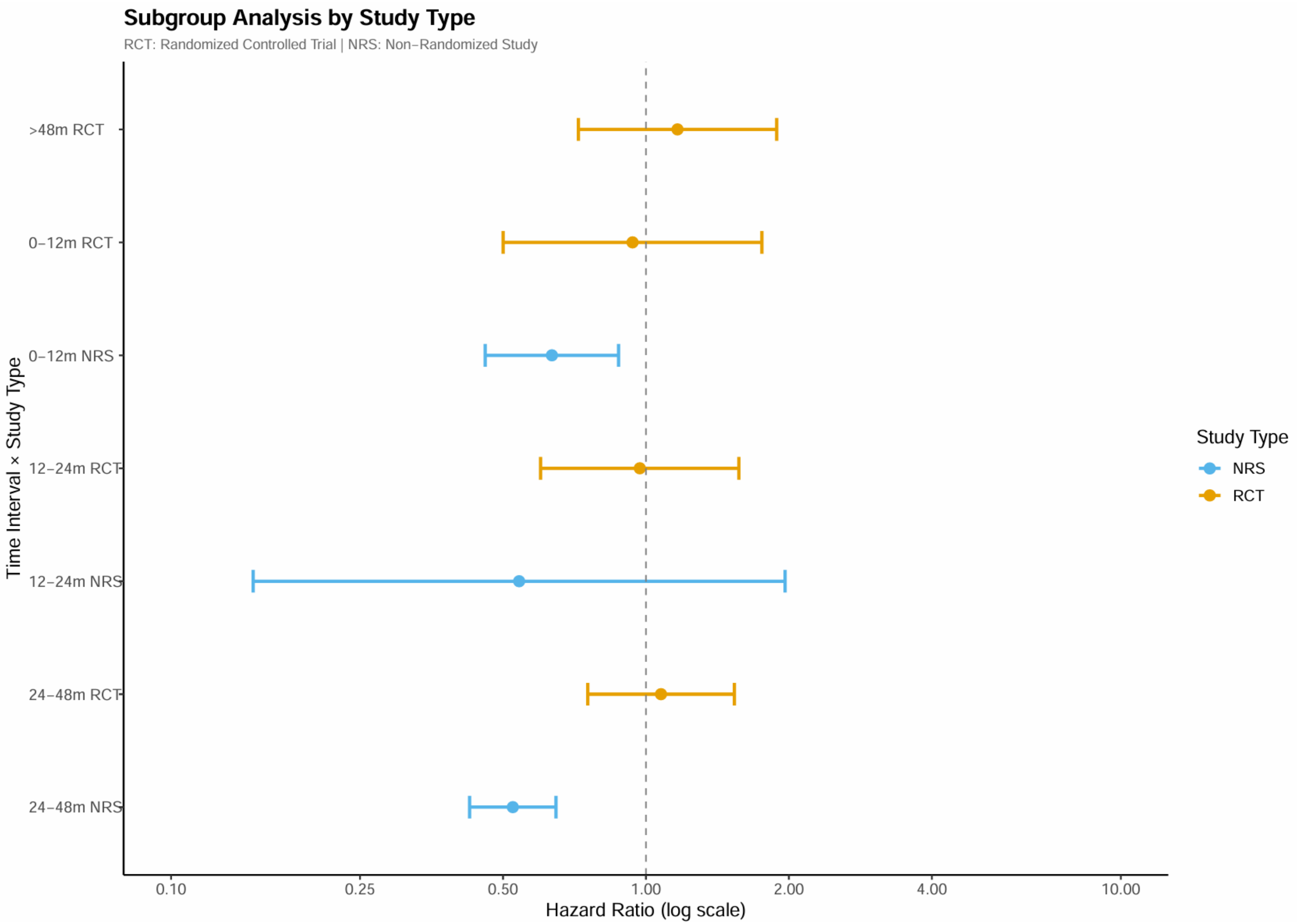

#### 6. Results of Sensitivity Analyses

##### 6.1 Leave-one-out sensitivity analysis of hazard ratios

Figure S5. Leave-one-out sensitivity analysis for cardiovascular endpoints

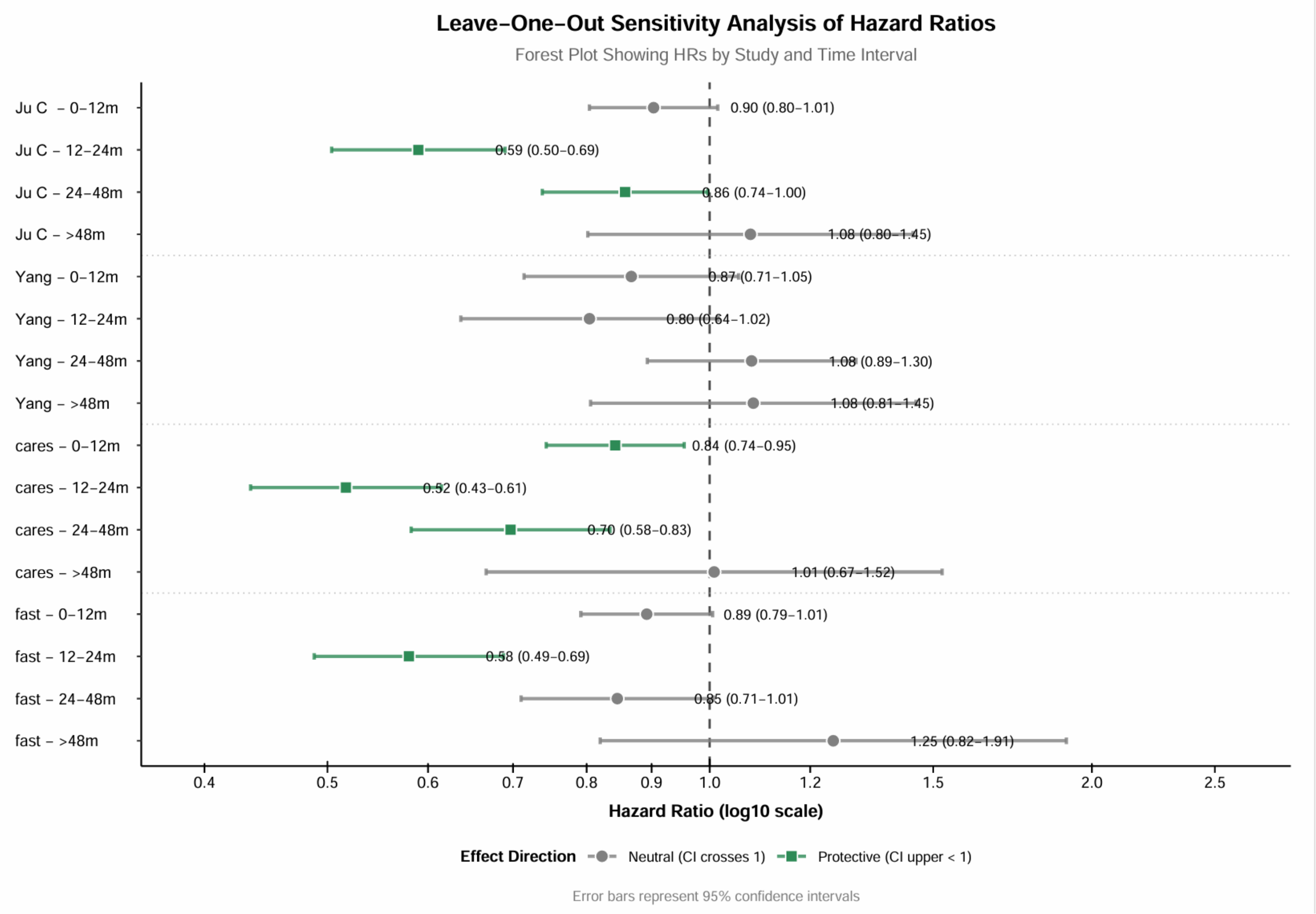

Figure S6. Leave-one-out sensitivity analysis for all-cause mortality

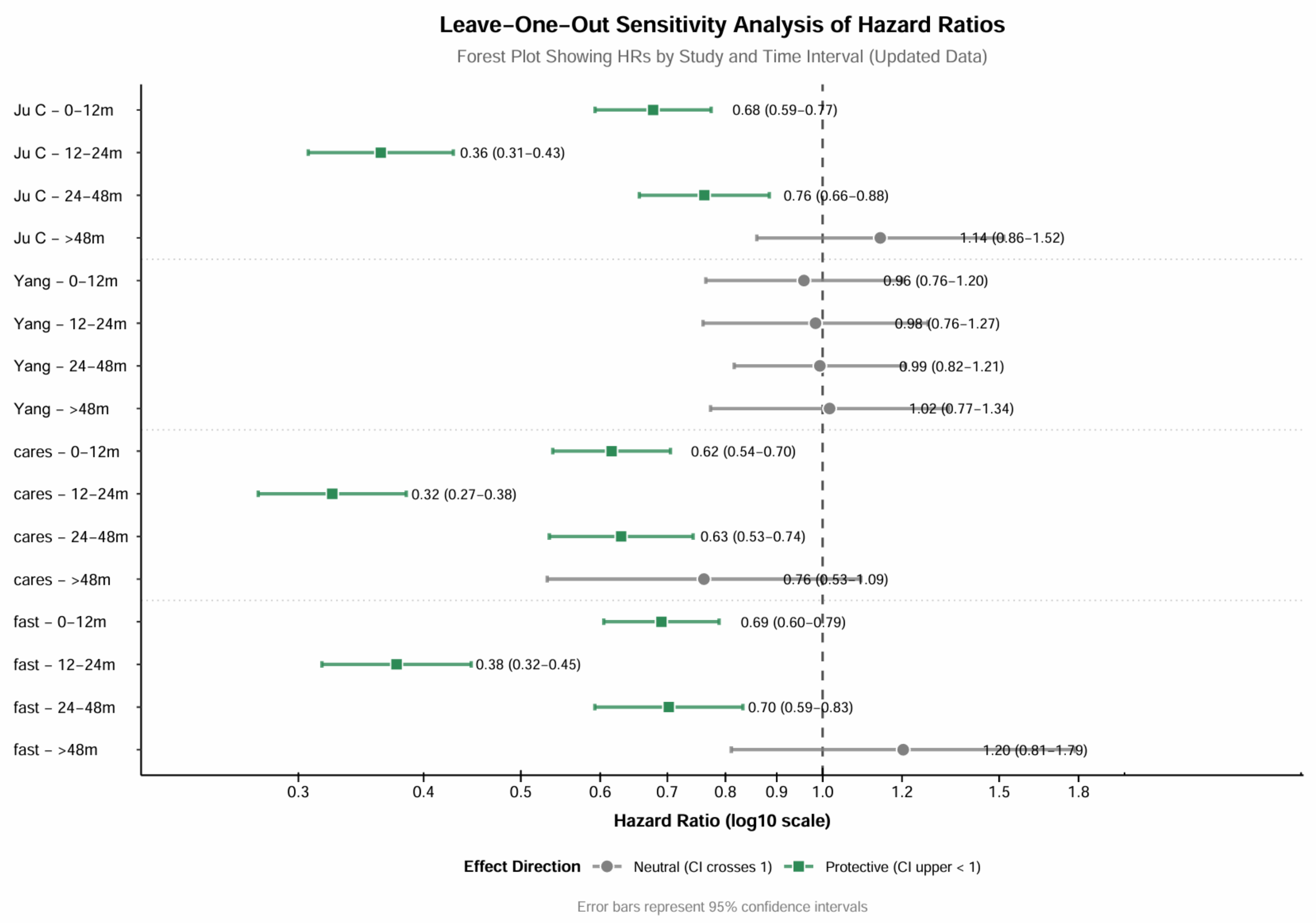

6.2 Bayesian Sensitivity Analysis

Figure S7. Bayesian analysis for cardiovascular endpoints

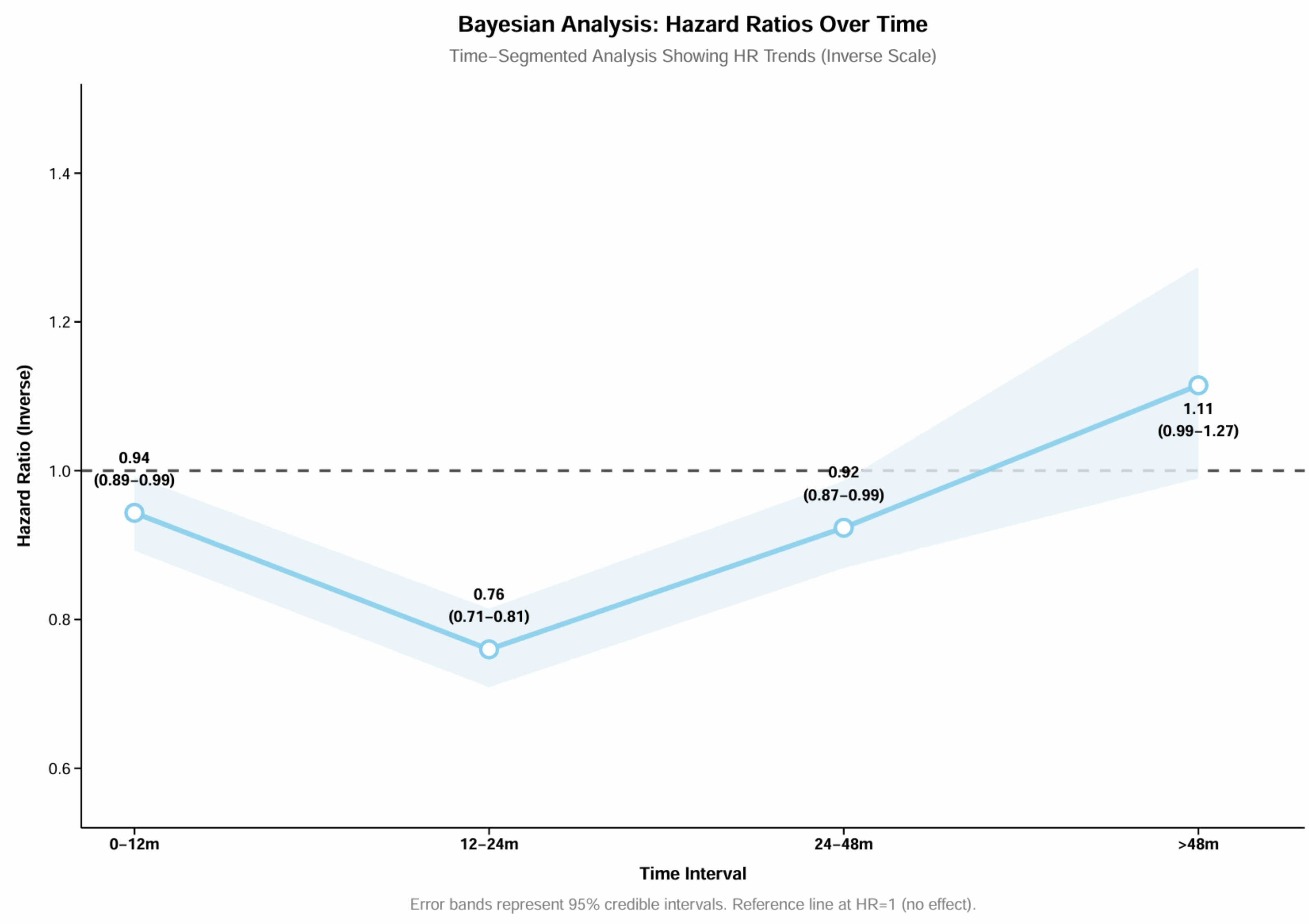

Figure S8. Bayesian analysis for all-cause mortality

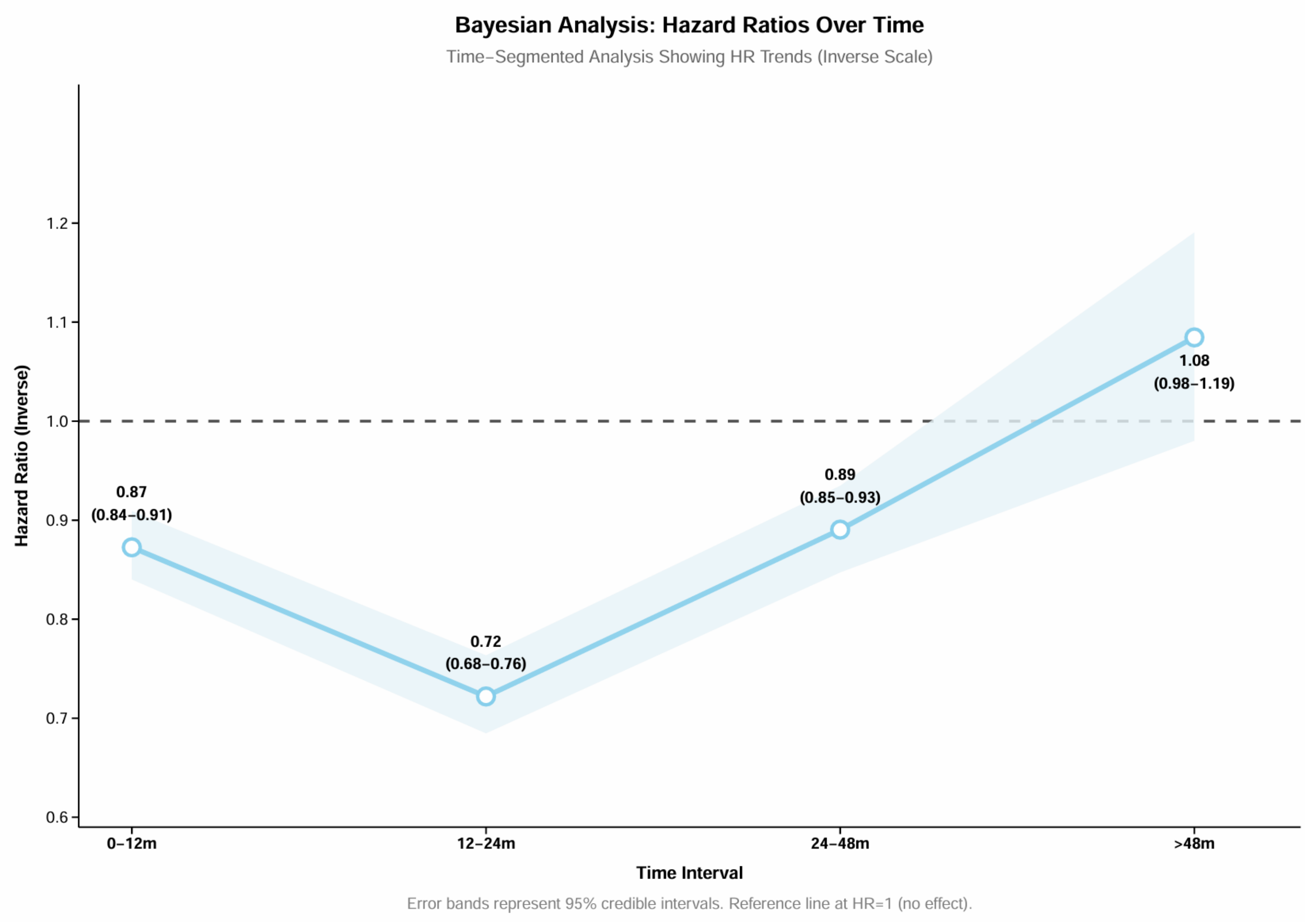

Figure S9. Two-stage meta-analysis for cardiovascular endpoints

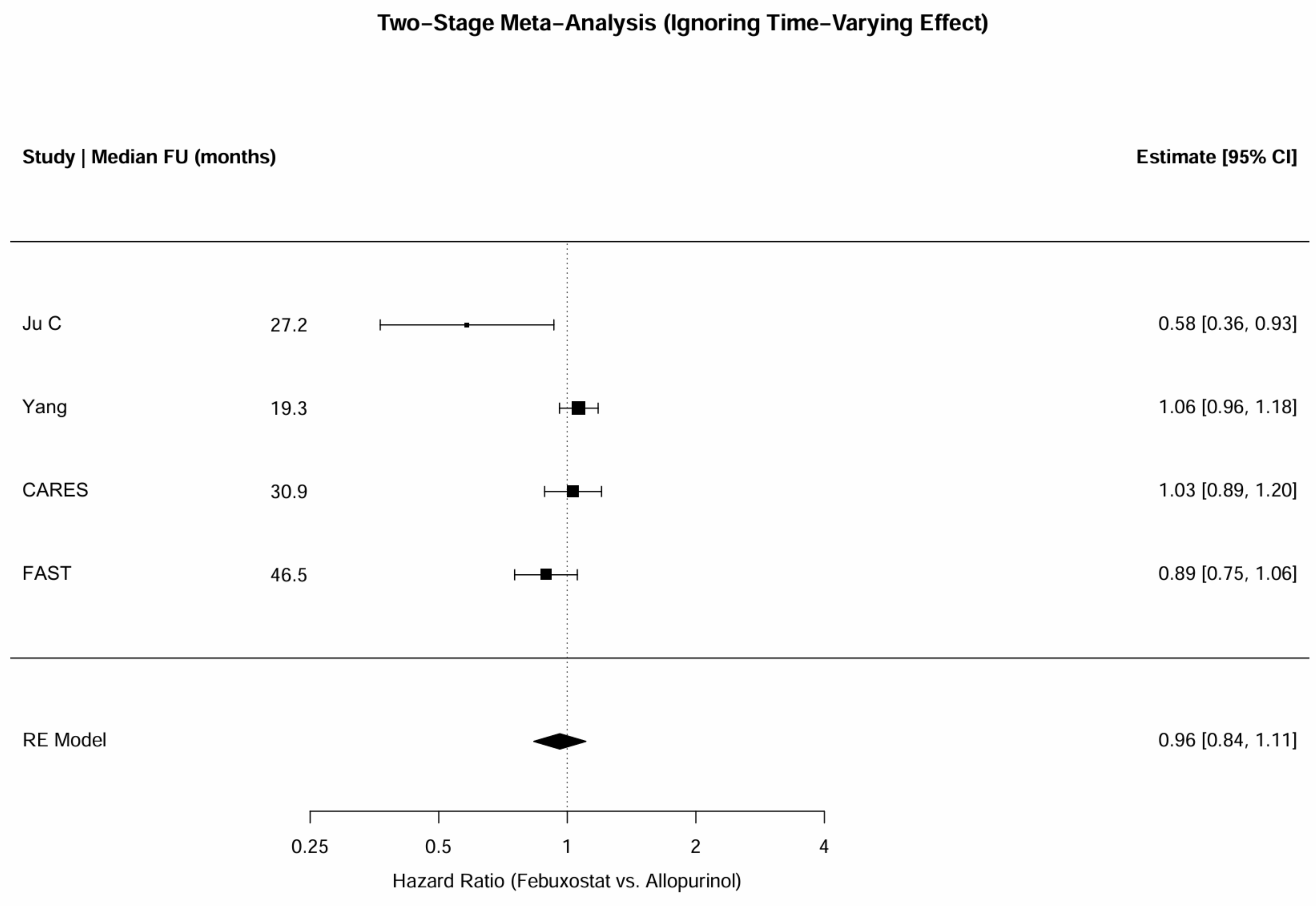

Figure S10. Two-stage meta-analysis for all-cause mortality

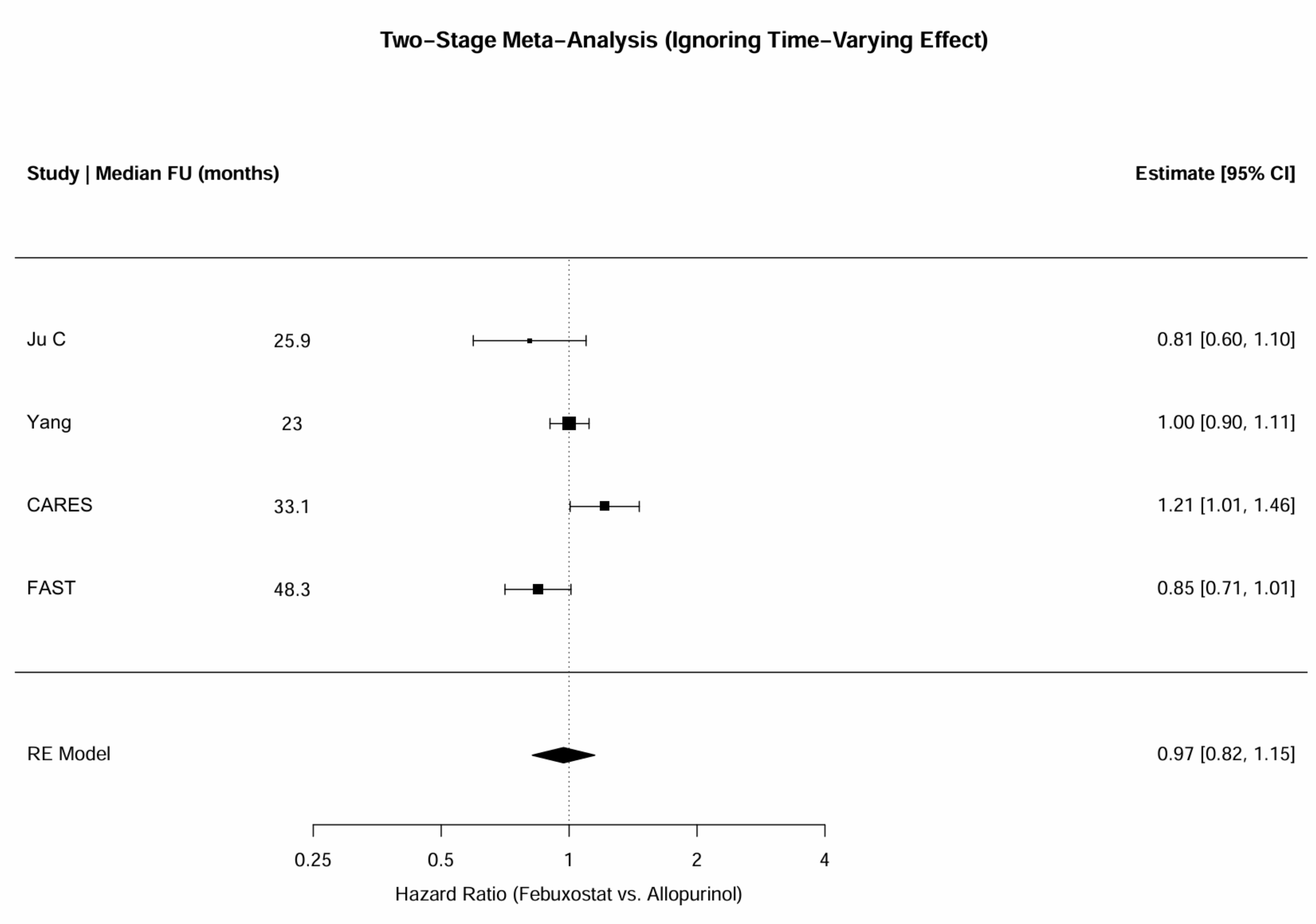

#### 7. Results of Publication Bias Assessment

Figure S11. Publication bias assessment for cardiovascular endpoints

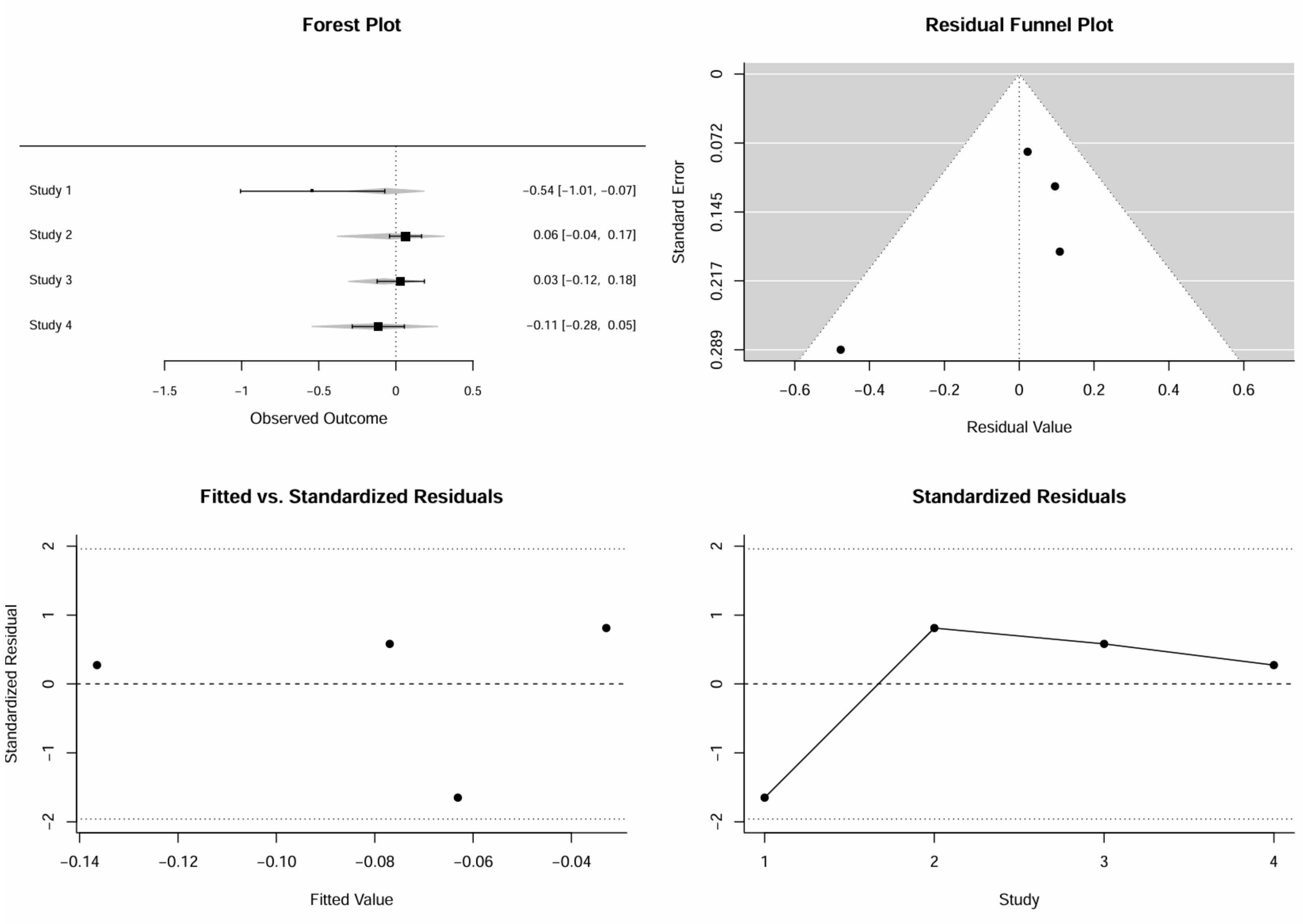

Figure S12. Publication bias assessment for all-cause mortality

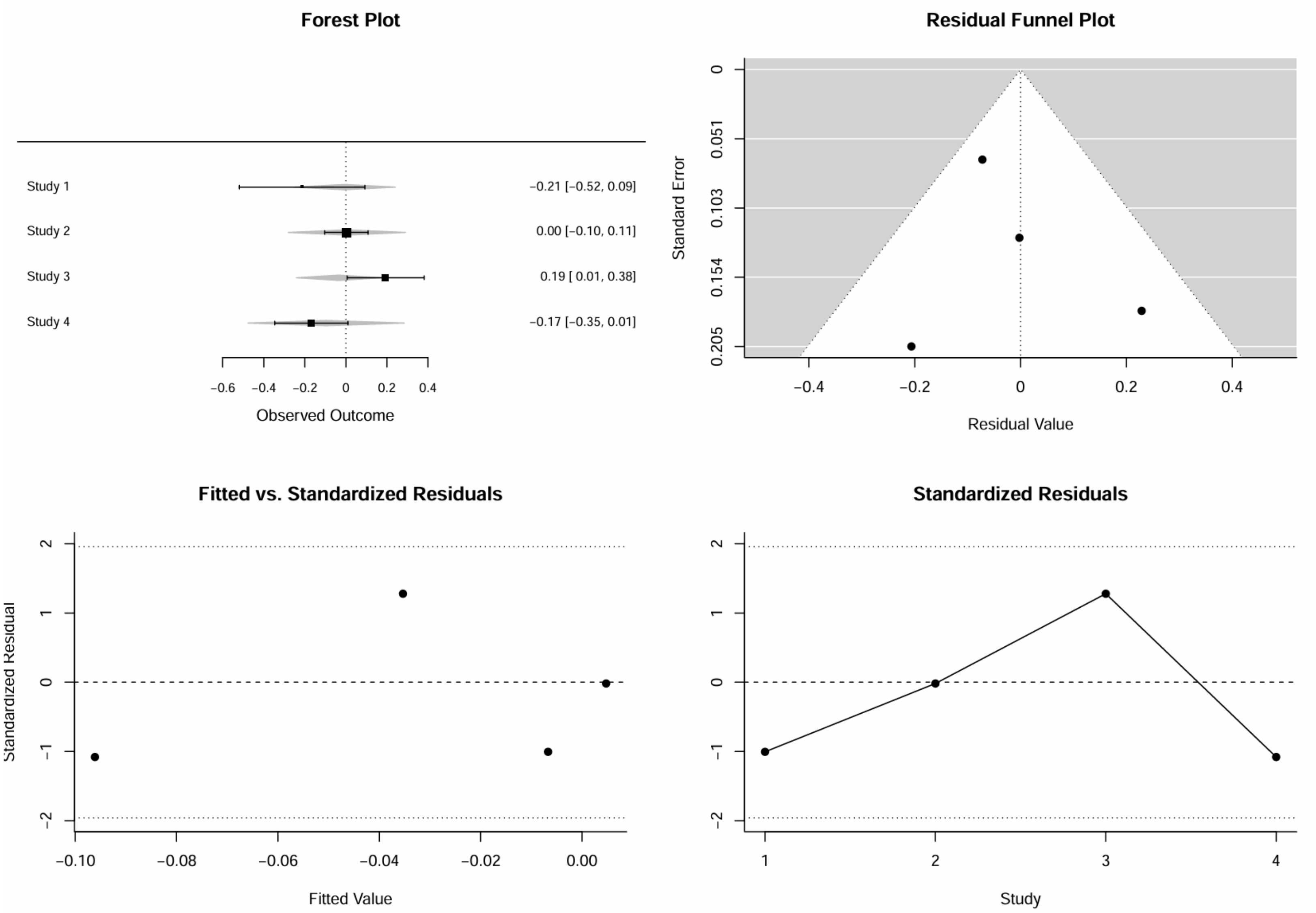

8. Kaplan-Meier Curves of Included Individual Study

Figure S13. KM curves of individual study for cardiovascular endpoints

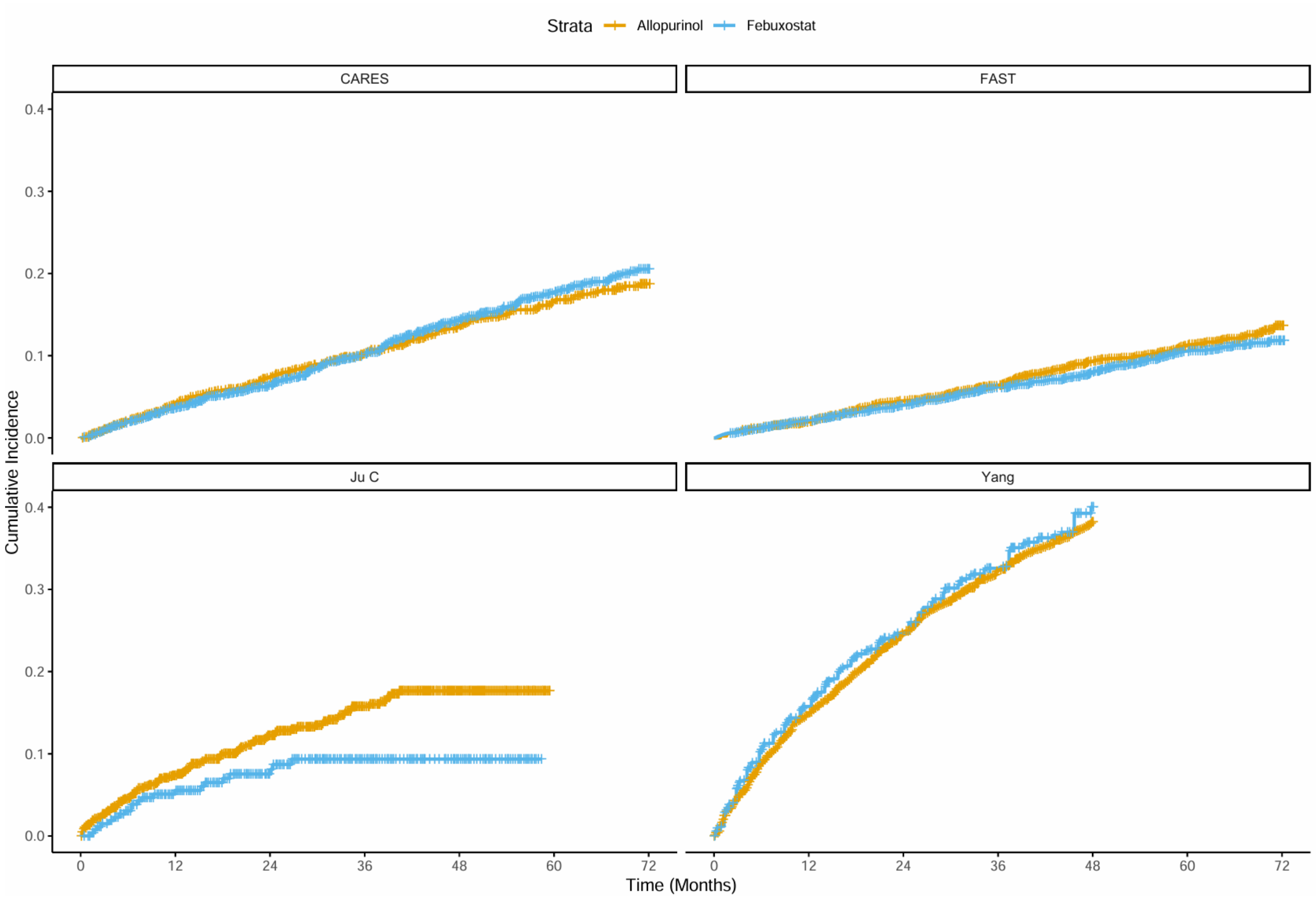

Figure S14. KM curves of individual study for all-cause mortality

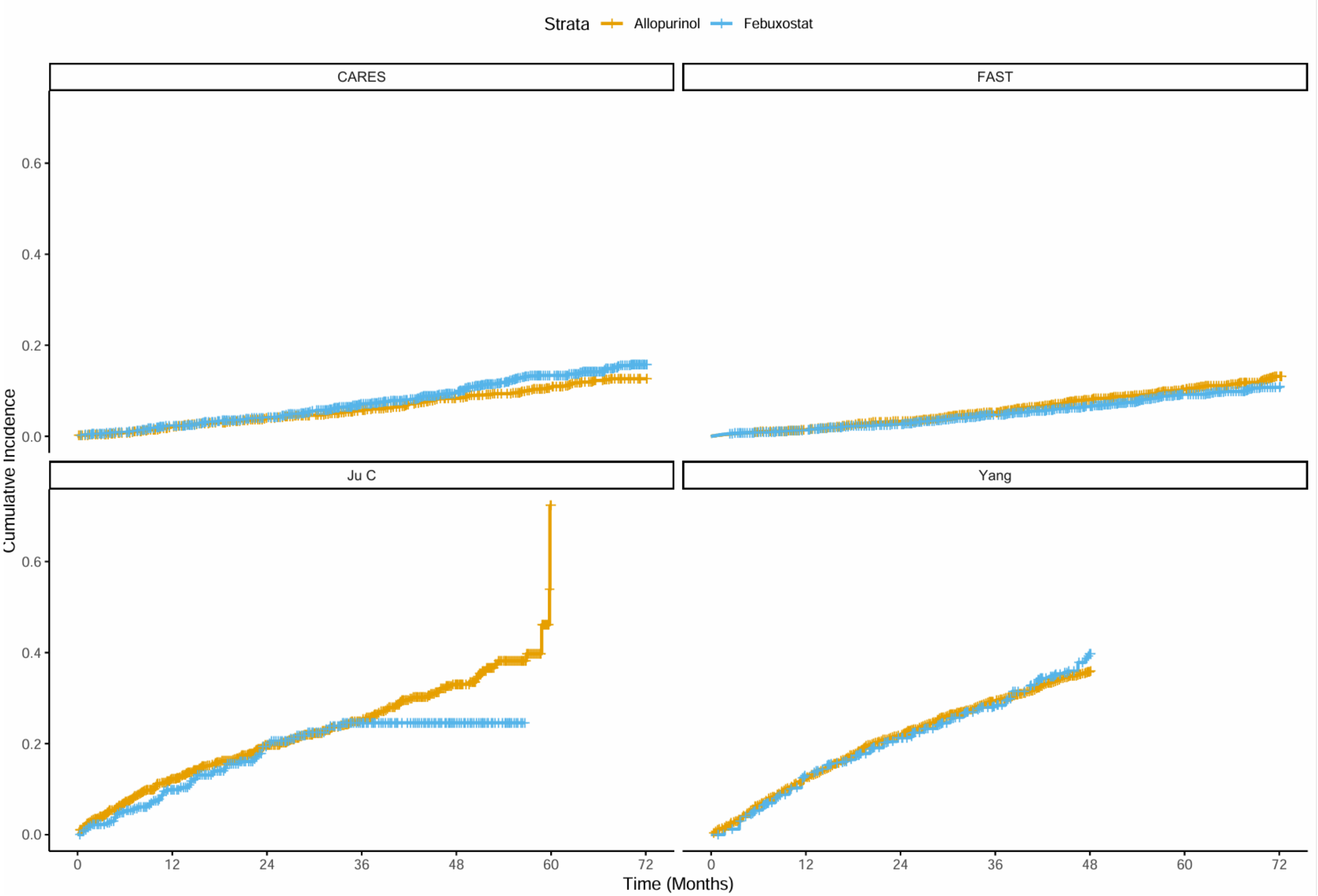

#### 9. Results of Additional Outcome (Cardiovascular Mortality)

Figure S15. Kaplan-Meier curve for cardiovascular mortality

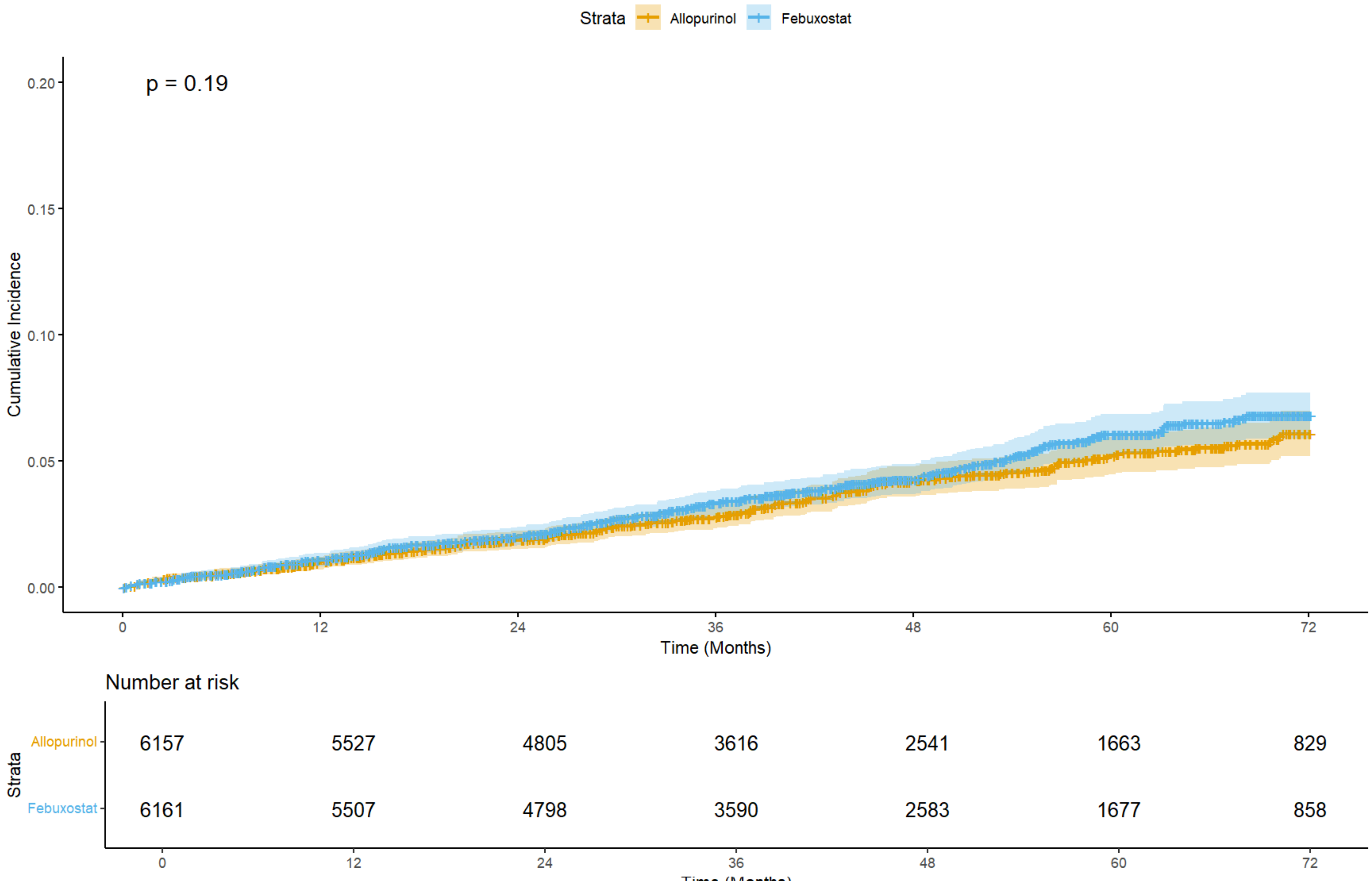

Figure S16. Kaplan-Meier curves for cardiovascular mortality by study

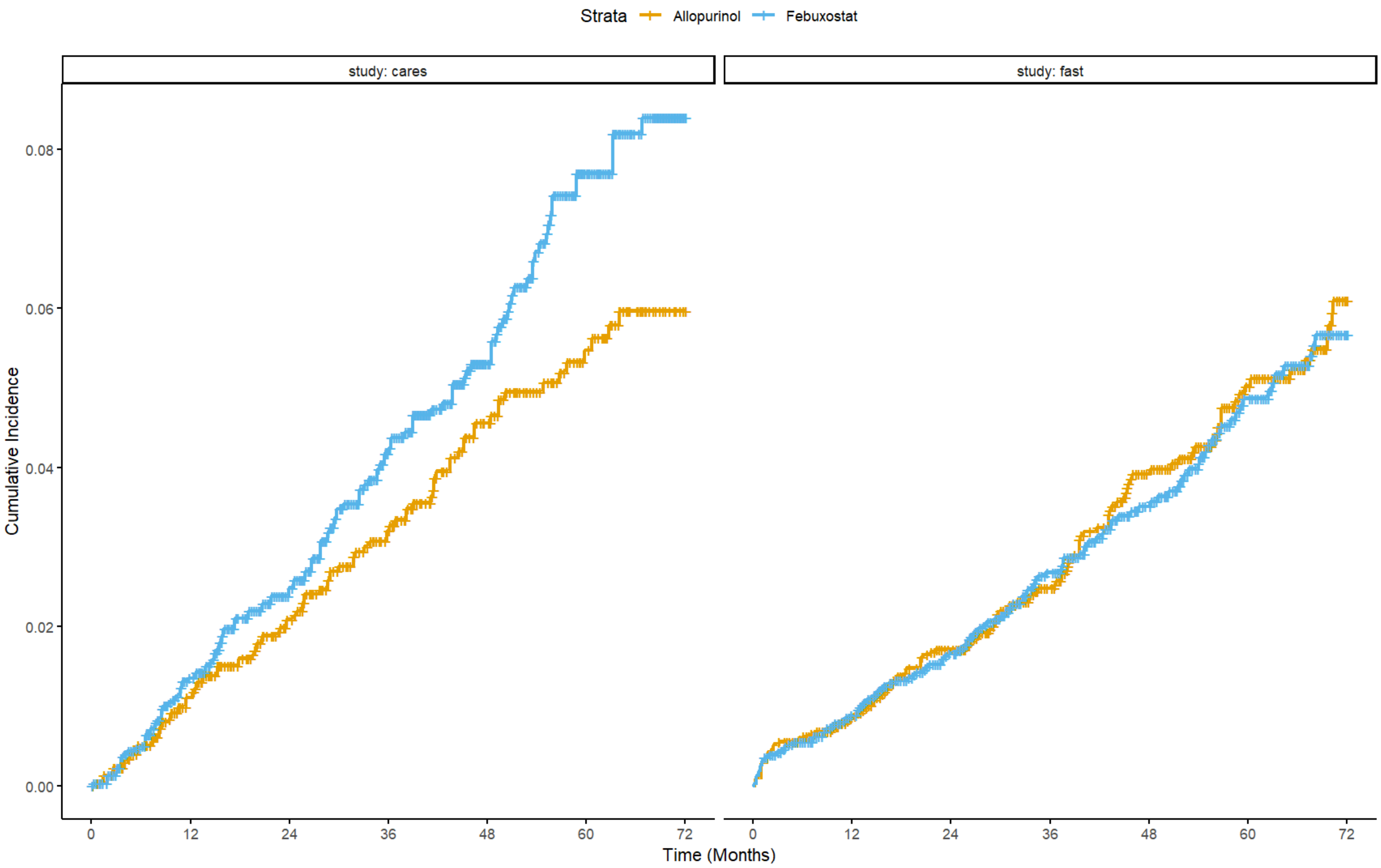

Figure S16. Result of main analysis for cardiovascular mortality

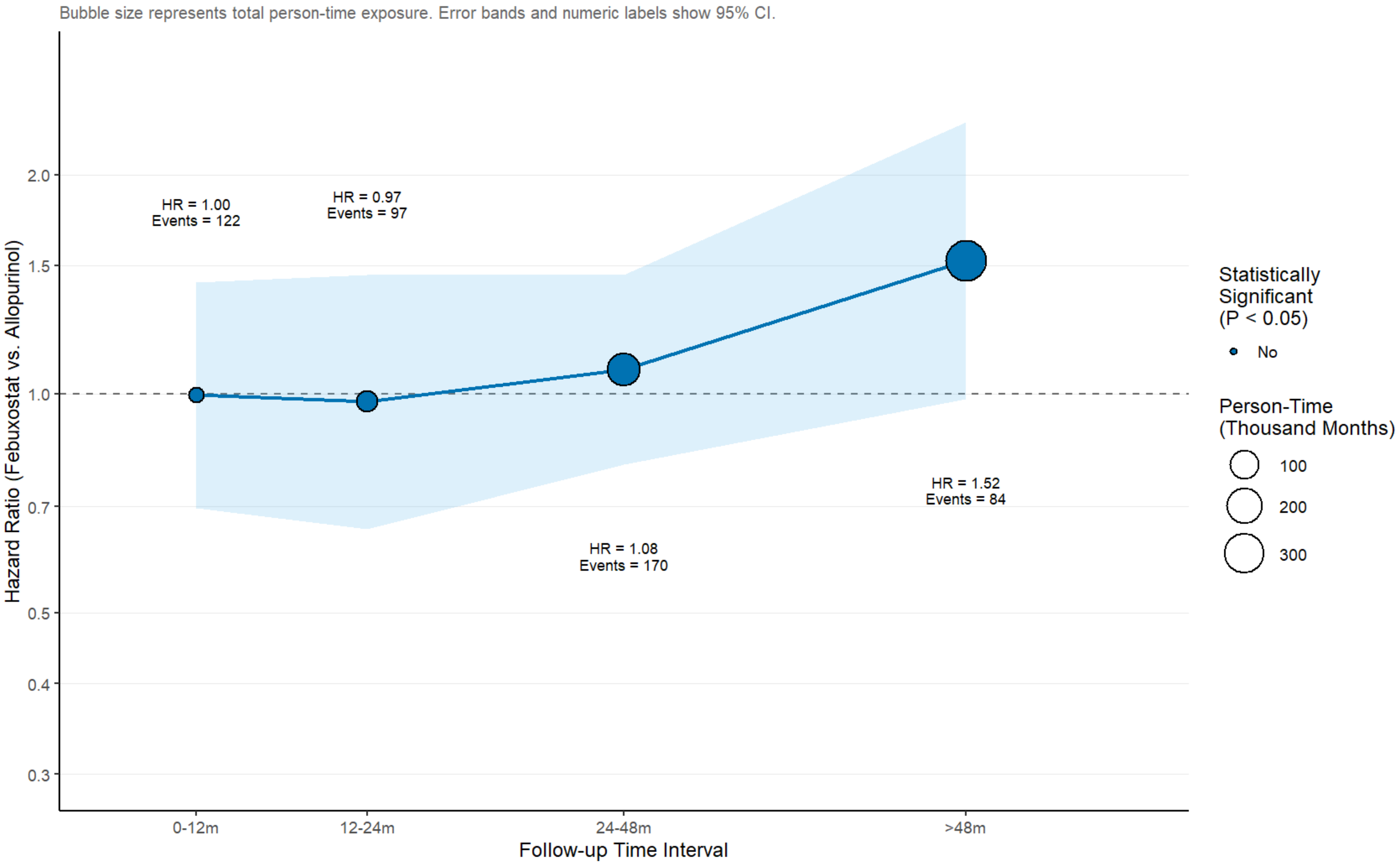
